# Resistance Training Frequency and Muscle Hypertrophy: A Systematic Review and Meta-Analysis of Volume-Equated Trials

**DOI:** 10.64898/2026.10.07.26365015

**Authors:** Anthony Gordon Boutagy, Mark McKean

## Abstract

Weekly training volume is the primary driver of resistance-training hypertrophy, but the independent role of frequency can be isolated only when weekly volume is equated. We quantified the effect of training frequency on hypertrophy across one to three volume-equated sessions per muscle per week. Following PRISMA 2020, five databases were searched to October 2026 for trials of at least six weeks that equated weekly volume between per-muscle frequencies and measured muscle size. One change-score standardized mean difference per trial (Hedges g; negative favoring higher frequency) was pooled with a random-effects model, and certainty was rated with GRADE. Sixteen trials (380 participants) were pooled. The frequency effect was trivial and non-significant (g = −0.06; 95% CI −0.24 to 0.13; I² = 0%; prediction interval −0.26 to 0.14); the interval admits at most a small effect favoring the higher frequency. Contrast-specific estimates were non-significant, trivial for one versus two and two versus three sessions and small for one versus three (g = −0.23; 95% CI −0.57 to 0.10), and no moderator was significant, although the moderator analyses could detect only large effects. Certainty was low to very low. Within this range, the data lack the precision to distinguish one, two or three sessions at a fixed weekly volume. Most trials measured muscle thickness, which does not track whole-muscle growth reliably.

## 1. Introduction

Resistance training increases muscle size through the repeated imposition of mechanical loading, and the magnitude of that increase is shaped by how the training variables are arranged. Among these variables, weekly volume, usually expressed as the number of hard sets performed per muscle per week, has the strongest and most consistent dose-response relationship with hypertrophy (Schoenfeld et al., 2017). Frequency, defined here as the number of times a given muscle is trained per week, is more ambiguous. It is frequently presented as an independent growth stimulus, on the reasoning that more frequent sessions sustain a higher time-averaged rate of muscle protein synthesis (Dankel et al., 2017). The difficulty is that frequency and volume are entangled: training a muscle more often, with sets per session held constant, raises weekly volume, so any apparent frequency effect may simply be a volume effect in disguise.

The only way to isolate frequency is to hold weekly volume constant between conditions, so that a muscle trained once per week performs the same weekly sets as a muscle trained two or three times per week, with the per-session set count adjusted accordingly. Three earlier meta-analyses bear on the question. Schoenfeld et al. (2016), pooling seven volume-equated per-muscle comparisons (200 participants, mostly untrained and mostly with indirect measures), reported an advantage for training a muscle twice rather than once per week (effect-size difference 0.19, 95% CI 0.11 to 0.28). Their update, with 13 volume-equated studies and a different effect-size model, revised this to a trivial and non-significant difference (0.07, 95% CI −0.02 to 0.17; Schoenfeld et al., 2019). Grgic et al. (2018) addressed strength rather than hypertrophy and found that the frequency effect on strength was no longer significant once volume was matched.

More recently, Pelland et al. (2026) modeled the weekly volume dose-response across a large corpus of trials and found the frequency coefficient for hypertrophy inconsistent across analyses and compatible with negligible effects, while their analysis restricted to the fifteen studies with direct within-study frequency comparisons did not indicate an effect of frequency; Tao et al. (2026) reported that frequency predicts strength but not muscle mass in volume-equated comparisons.

What these syntheses share is volume equation; what they leave open is a focused estimate for the frequency range that practice most often uses. Schoenfeld et al. (2019) and Tao et al. (2026) pooled frequencies above three sessions per muscle per week, so their estimates average across schedules that few programs adopt, and both treated several sites or time points from one trial as separate effects. The present review therefore (i) restricts to per-muscle, volume-equated hypertrophy trials within the ecologically common range of one to three sessions per week; (ii) derives one change-score standardized mean difference per trial directly from the trial reports, with transparent handling of imputed dispersion and a stated smallest effect size of interest; (iii) applies small-sample random-effects methods, reports what its moderator analyses could and could not detect, and rates certainty with GRADE; and (iv) frames frequency explicitly as a distribution lever for weekly volume rather than as a competing stimulus.

## 2. Materials and Methods

### 2.1. Protocol and reporting

The review was conducted and reported in accordance with the PRISMA 2020 statement (Page et al., 2021), with specific attention to the sport and exercise science guidance (PERSiST; Ardern et al., 2022). The review was not prospectively registered. An Open Science Framework (OSF) project (https://osf.io/ebrcx) describing the review question, eligibility criteria, primary and secondary outcomes (change in muscle size; moderation by intervention duration and training volume), and the risk-of-bias and certainty frameworks was created on 24 June 2026, after the original searches (April 2026) and data extraction; it is a summary of the planned review rather than a full protocol, and it does not specify the effect-size formulas or the sensitivity analyses.

The searches reported here were run on 6 October 2026 and replace the April 2026 searches, for which complete screening records were not retained. The extracted data, the screening log for every record and the analysis code are deposited in the same OSF project, so that every effect size and pooled estimate can be reproduced and re-analyzed under alternative assumptions (for example, different pre-post or within-trial site correlations). The review was not registered on PROSPERO because registration is restricted to reviews that have not yet completed data extraction. This version differs from the first preprint (medRxiv, 10 October 2026) in one input: the effect size for the within-subject trial of Neves et al. (2022) is now taken from its sets-equated condition rather than its volume-load-equated condition, for the reasons given in Section 2.5, after a reader of the preprint questioned the choice; the first-version estimate is retained as a sensitivity analysis (Section 3.3). Version 3 corrects two reporting errors (Supplementary Methods S7; Section 4.6), revises wording in the Abstract, Sections 4.3, 4.4 and 4.8 and the Conclusions so that non-significant results are not read as evidence of equivalence, and adds to Section 4.5 and Supplementary Note S14 on swelling across sessions and maintenance intervals; no estimate, figure or table changed. Version 4 corrects the volume-load difference between limbs in the sets-equated condition of Neves et al. (2022) from about 17% to 19.5% in Sections 2.5, 4.4 and 4.6 and Supplementary Methods S9 (164,894 against 137,986 kg in the trial’s Results; the authors’ Conclusion gives 17%), gives the Brigatto et al. (2019) difference in Section 4.6 to the same precision (16.3%), and corrects the description of Mpampoulis et al. (2024) in Section 4.5 and Supplementary Note S14: the 94% reported by the authors is the share of post-training cross-sectional area kept, not of the gain, and one session every fourteen days lost about half the mean gain of the whole training cohort (the subgroup’s own gain was not reported); no estimate, figure or table changed. The declaration of generative AI use adds the authors’ confirmation that they have verified the analyses and take responsibility for the manuscript and the deposited materials, as in the version submitted to Communications in Kinesiology.

Analyses not named in the OSF project description are identified as such where they are described: the coding of measurement timing, the moderator analyses for repetitions and relative load, the training-status subgroup, the alternative confidence-interval conventions, the sensitivity analyses for the pre-post and within-trial correlations, the blinded-assessment and direct-measure one-versus-three subsets, the alternative dispersion sources for Brigatto et al. (2022), the analyses removing the four added trials and Gentil et al. (2018), and the sensitivity inclusion of Tavares et al. (2017).

### 2.2. Eligibility criteria

Trials were eligible if they (i) used a longitudinal resistance training intervention of at least six weeks; (ii) compared, for the measured muscle, training frequencies of one, two or three sessions per week, so that the eligible contrasts were one versus two, one versus three and two versus three, and a comparison in which the measured muscle was trained more than three times per week in any arm was ineligible; (iii) assigned the frequencies by the investigators rather than by self-selection or attendance; (iv) equated total weekly volume between conditions, with the per-session set count adjusted to hold weekly sets, repetitions and load constant; and (v) reported a direct or indirect measure of muscle size (ultrasound or magnetic resonance imaging muscle thickness or cross-sectional area, or an estimate of lean mass by dual-energy x-ray absorptiometry or anthropometry, including skinfold-derived lean mass).

Bioelectrical impedance was not accepted as a measure of muscle size; one trial was excluded on that ground (Cardoso et al., 2024), and its split arm trained each muscle 1.5 times per week on average, outside the eligible contrasts. Designs that fixed per-session sets and therefore allowed weekly volume to differ between frequencies were excluded, because they confound frequency with volume. Where a trial compared frequencies for some measured muscles and not others, only the muscles trained at an eligible contrast were used (Section 2.4).

Throughout, hypertrophy is used as the umbrella construct, defined as an increase in muscle size, and its operationalizations are distinguished: muscle size denotes direct site-specific measures (ultrasound or magnetic resonance imaging muscle thickness or cross-sectional area), whereas lean or muscle mass denotes indirect whole-body or regional estimates (dual-energy x-ray absorptiometry or skinfold-derived). Because the indirect measures are less sensitive to localized growth, this distinction is carried through the analysis and its limitations.

These criteria were chosen to isolate the independent influence of per-muscle training frequency on hypertrophy under conditions that closely mirror common practice: ecologically typical frequencies (one to three sessions per week), strict per-muscle weekly volume-equation, and direct or clinically relevant hypertrophy outcomes. This focus avoids the volume-frequency confounding that characterizes much of the broader literature and positions the pooled estimate as a direct test of whether redistributing a given weekly volume across the week alters hypertrophic adaptations. One trial (Tavares et al., 2017) compared one with two sessions per week with sets equated (four sets of each exercise once weekly against two sets twice weekly) and volume-load described by its authors as equalized, but did so during an eight-week reduced-training phase, at about half the preceding volume-load, that followed an eight-week standardized program, so that neither arm gained muscle size; because the quantity it measures is retention under a reduced dose rather than growth, it was analyzed in a sensitivity analysis rather than pooled (Section 3.3).

### 2.3. Information sources and selection

Five databases were searched on 6 October 2026 with no date limit: MEDLINE through PubMed, Scopus, Web of Science Core Collection, SPORTDiscus through EBSCO, and the Cochrane Central Register of Controlled Trials (CENTRAL) through Ovid EBM Reviews. Each strategy combined three groups of terms with AND: resistance or strength training; training frequency (for example “training frequency”, “sessions per week”, “once per week”, “split routine”, “full body”); and muscle size (for example “hypertrophy”, “muscle thickness”, “cross-sectional area”, “lean mass”). Records were limited to English. The full strategies, with the database-specific syntax and limits, are given in Supplementary Methods S1, and the number of records from each database in Supplementary Table S3. The reference lists of seven frequency reviews (Cuthbert et al., 2021; Grgic et al., 2018; Kneffel et al., 2021; Ramos-Campo et al., 2024; Schoenfeld et al., 2016, 2019; Tao et al., 2026) were then searched by hand. For one of these (Kneffel et al., 2021) only the reference list could be read, not the table of included studies. The dose-response meta-regression by Pelland et al. (2026) was used to frame the question but not as a source of trials, because its included set is not restricted to volume-equated designs.

Records were deduplicated by DOI, then by normalized title, then by near-identical title in the same publication year. Titles and abstracts were screened against the eligibility criteria, and every record not excluded at that stage was assessed against the full criteria, on the full text where the abstract left any criterion open and on the abstract alone where it stated the failing design feature explicitly (for example, a four-session arm or the same per-session prescription in every arm). Every decision, with its reason and the basis on which it was made, is recorded in the deposited screening log; the exclusions at eligibility assessment are listed with reasons in Supplementary Table S2. One trial (Tavares et al., 2017) met the criteria but measured retention during a reduced-training phase and was handled as described in Section 2.2.

For the original twelve trials, data extraction was performed primarily by one reviewer; for the four trials added to the pool in October 2026, data were extracted with the generative AI tool declared in the end matter and checked against the trial reports by the authors. To assess accuracy, a second reviewer independently re-screened and re-extracted a random and targeted subsample of seven trials of the original set: five randomly selected trials and the two trials flagged at screening in the original set for absent dispersion data (Gentil et al., 2018, before its participant-level data were retrieved; Schoenfeld et al., 2015), with the random seeds documented; Candow and Burke (2007), whose dispersion was also imputed, fell outside the random draw.

The second reviewer also re-extracted the four trials added to the pool in the October 2026 update (Gentil et al., 2018; Yue et al., 2018; Prieto González et al., 2020; Calzavara & Barroso, 2026), so that 10 of the 16 pooled trials were checked. Percentage agreement was calculated for eligibility decisions and categorical extraction fields; Cohen’s kappa is undefined here because every re-screened trial was an include, so expected agreement equals observed agreement.

All sixteen pooled effect sizes were also recomputed from the trial reports with the same tool, under the rules in Section 2.4, in code separate from the authors’ own; for the four added trials this checks the computation but not the extraction, which the same tool performed. The tool also ran the October 2026 database searches through institutional library access, performed the title and abstract screening, and assessed records against the eligibility criteria; every decision at eligibility assessment and every extracted value was checked by the authors, who confirmed all 79 eligibility decisions, and the authors also checked a random sample of 100 of the 1,823 title and abstract exclusions other than reviews (seed documented) and confirmed all 100.

The check verified eligibility decisions and extracted fields; it did not substitute for dual-independent review.

### 2.4. Data extraction

For each trial we extracted, per group and per measured site, the pre and post means and standard deviations, the sample size, the per-muscle frequency contrast, the intervention duration, the training status of the sample, and the hypertrophy measurement method. We also extracted, from each trial’s Methods, the interval between the final training session and the post-intervention muscle-size measurement, and whether any index of muscle edema (for example ultrasound echo intensity) was reported. The interval was coded as 48 hours or longer where it was stated or clearly implied by the measurement procedure, and as not reported otherwise. This coding was added after the primary analysis, in response to the concern that transient post-exercise swelling could inflate muscle-size measures taken soon after training (Damas et al., 2016), and the corresponding sensitivity analysis is reported in Section 3.3.

Frequency was coded per measured muscle from each trial’s exercise schedule; where a split routine trained the measured muscle as a prime mover on more than one day, the contrast was coded from the sessions in which the muscle was trained directly (Schoenfeld et al., 2015; Brigatto et al., 2019; Lasevicius et al., 2019), and the per-session and weekly set counts used as moderators follow the same rule (Supplementary Table S4). In Yue et al. (2018) the lower body was trained only in the second of two routines, once weekly in one arm and twice weekly in the other, whereas the upper-body muscles were also worked in the first routine and so were trained twice versus four times per week; only the vastus medialis site, a one-versus-two contrast, was used.

One rule was applied to dispersion. Where a trial reported change-score dispersion directly, those values were used: change-score standard deviations for Neves et al. (2022), Thomas and Burns (2016) and Yue et al. (2018); standard deviations recovered from the 95% confidence intervals of the mean change for Grzyb et al. (2022), where the interval printed for the knee-extensor change in the three-session group (0.31, 0.09 to 0.12 cm) does not contain its point estimate and was reconstructed symmetrically from its lower bound (0.09 to 0.53 cm); and the trial-reported between-group standardized differences of the changes for Brigatto et al. (2022), which are consistent with that trial’s group-by-time tests at three of its four sites, whereas the standard deviations implied by its printed confidence intervals of the mean change are not (Supplementary Table S5 gives the composite under each source). Pina et al. (2020) reported covariate-adjusted mean changes with confidence intervals; because the two groups’ changes were equal, its effect size is 0.00 under either source.

Gentil et al. (2018) presents muscle thickness only as an individual-values figure; its effect size was computed from the participant-level data published with the trial, which give each participant’s change directly. Where only pre and post standard deviations were reported, the change-score standard deviation was derived with an assumed pre-post correlation (Section 2.5).

Two trials did not report usable dispersion for the post-intervention values: Schoenfeld et al. (2015) present means with standard deviations in figures only, and Candow and Burke (2007) report baseline standard errors in a table and post-intervention values in a figure, with the percentage change stated differently in the Abstract (2.2%) and the Results (2.9% and 3.0%); for both, the standard deviations were read from the figures and the derived change-score standard deviations are treated as imputed, flagged, and tested in sensitivity analysis.

### 2.5. Effect size and synthesis

#### Effect measure

The effect measure was the change-score standardized mean difference (Hedges g), computed as the between-group difference in mean change divided by the pooled change-score standard deviation, with the small-sample correction applied (Hedges, 1981). The sign convention was set so that a negative g favors the higher-frequency condition. A smallest effect size of interest was set at |g| = 0.2, the conventional boundary between a trivial and a small standardized effect, so that pooled point estimates below this bound are interpreted as trivial in magnitude, and only intervals contained within it as excluding even a small effect. Where only pre and post standard deviations were reported, change-score standard deviations were derived from them using a pre-post correlation of r = 0.7, a value within the range commonly assumed for change-score analyses when trial-reported test-retest correlations are unavailable, with sensitivity analyses at r = 0.5 and r = 0.9. The assumed correlation is the denominator of every such effect size, which scales by the square root of 0.6 divided by 2(1 - r), so the sensitivity analysis reports the pooled estimate, its heterogeneity and the most affected trials at each value.

#### Multi-site trials

Trials reporting multiple sites were aggregated to a single composite effect using the dependent-effects formula of Borenstein et al. (2009), assuming a within-study correlation among sites of 0.6; the correlation enters the composite through its sampling variance, and therefore through the trial’s weight in the pool, so the pooled estimate was recomputed at correlations of 0.3, 0.8 and 1.0.

#### Pooling and confidence intervals

Effects were pooled with a random-effects model with the between-trial variance estimated by restricted maximum likelihood; where that estimate is zero, as it was in every model here, the estimate coincides with the DerSimonian and Laird (1986) estimate. Confidence intervals are reported with one convention throughout: the Wald interval from the inverse-variance weights. The Hartung-Knapp-Sidik-Jonkman (HKSJ) adjustment, which is often recommended for small meta-analyses, scales the standard error by the observed dispersion of the trials around the pooled estimate; when the trials agree more closely than chance would produce, as here, the unmodified adjustment narrows the interval below the Wald interval, and the standard remedy is to truncate the scaling factor at one. The truncated HKSJ interval is reported alongside the Wald interval for the overall estimate; at the contrast level, with five or six trials, it is wider than the Wald interval, so it is given in the Supplementary Material (Table S6).

#### Heterogeneity and sensitivity analyses

Heterogeneity was summarized with I² (Higgins et al., 2003) and a prediction interval. The stability of the pooled estimate was examined by leave-one-out analysis, by restricting to direct measures of muscle size, by removing trials with imputed variance, by removing the trials rated at high risk of bias, by restricting to trials whose final muscle-size measurement was taken 48 hours or more after the last training session, by training status, by removing the four trials added in the October 2026 update, and by adding Tavares et al. (2017) (Section 2.2).

#### Moderator analyses

To examine whether intervention length modified the effect, the change-score effect sizes were regressed on trial duration in weeks using a random-effects (method-of-moments) meta-regression, with the slope, its 95% confidence interval, and residual heterogeneity reported. The moderator analyses (duration, per-session and weekly volume, mean repetitions and relative load) were exploratory and interpretively secondary; with at most 16 trials per model and no correction for multiple testing, they are reported as hypothesis-generating rather than confirmatory, and for each regression the smallest slope detectable with 80% power at two-sided alpha of 0.05 (2.8 standard errors) is reported alongside the estimate. The moderator values and the counting rule are tabulated in Supplementary Table S4.

#### Within-subject trial and small-study effects

The within-subject trial (Neves et al., 2022) ran two conditions in separate groups of twelve trained men, each man training one leg once and the other three times per week for the same nine weekly sets of leg press. In the first condition both legs trained at their own repetition-maximum loads, so that the three-session leg accumulated 19.5% more volume load (164,894 against 137,986 kg; the authors’ Conclusion gives 17%); in the second the three-session leg was held to the once-weekly leg’s mean load per repetition, so that volume load was also equal, which the authors note meant its sets were not always maximal. The sets-equated condition was used, because it matches the prescription rule applied to every other trial in the pool (equal weekly sets, the same repetition-maximum prescription in each arm, and achieved volume load free to differ, as in Brigatto et al., 2019, and Calzavara & Barroso, 2026) and the coding of the same trial in the open corpus analyzed by the companion paper (Section 4.2); the volume-load-equated condition is reported as a sensitivity analysis (Section 3.3). Because the trial reported contralateral limbs without a within-limb correlation, its variance was computed with the independent-groups standardized-mean-difference formula, which widens its interval and discards the precision of the within-subject design. Small-study effects were examined with Egger’s regression using both a standard-error moderator and a sample-size-based (1/√n) precision term, with Neves et al. (2022) entered as 24 limbs. Analyses were performed in Python 3.11 (NumPy 1.26 and SciPy 1.11); the pooled and contrast estimates, τ², I², the duration meta-regression and Egger’s test were reproduced in the R package metafor as an independent cross-check, and both implementations are deposited on OSF.

### 2.6. Risk of bias and certainty

Risk of bias was appraised with the Cochrane RoB 2 tool (Sterne et al., 2019), and the certainty of evidence for each frequency contrast was rated using the GRADE approach (Guyatt et al., 2008), with the optimal information size computed for the smallest effect size of interest. Appraisal was performed by a single reviewer, supported where relevant by PEDro-scale criteria (de Morton, 2009). The principal sources of bias in this literature are small samples, short interventions, unblinded outcome assessment, per-protocol analysis after exclusions for non-adherence and, in two trials, incompletely reported dispersion. The second-reviewer check (Section 2.3) verified screening and extraction rather than re-appraising risk of bias, so the risk-of-bias judgments remain single-reviewer; this is reflected in the GRADE downgrades for risk of bias and imprecision.

## 3. Results

### 3.1. Study selection and included trials

The study selection process is summarized in Figure 1. The five databases returned 3,605 records, of which 1,935 remained after duplicates were removed. Title and abstract screening excluded 1,856, and 79 were assessed against the full criteria, of which 66 were excluded (Supplementary Table S2). Of the remaining 13 records, 11 were pooled trials, one was a second report of a pooled trial (Pina et al., 2020) and one was the reduced-training trial analyzed in sensitivity analysis only (Tavares et al., 2017). One pooled trial was published after the original April 2026 search (Calzavara & Barroso, 2026). Ten further reports were assessed from other sources: nine from the reference lists of the seven reviews and one, Brigatto et al. (2022), identified in the April 2026 search and not retrieved by the October 2026 strings. Five of these were included (Gentil et al., 2015; Grzyb et al., 2022; Brigatto et al., 2022; Yue et al., 2018; Prieto González et al., 2020).

**Figure 1.**
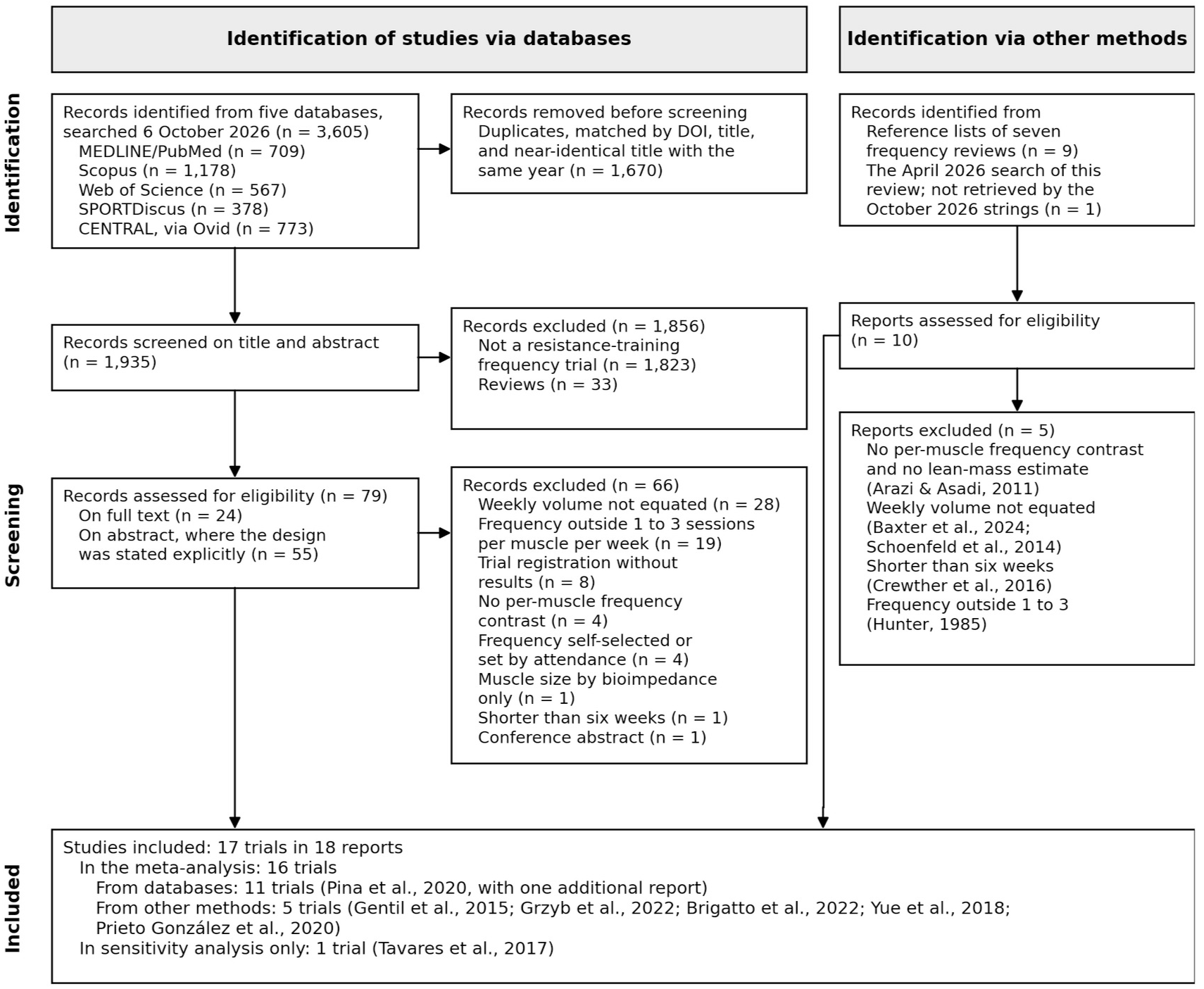
PRISMA 2020 flow diagram for study selection. Five databases were searched on 6 October 2026 (3,605 records; 1,935 aker duplicate removal); 79 records were assessed for eligibility and 10 further reports from other sources. Sixteen trials were pooled and one was analyzed in sensitivity analysis only. The reports excluded from other sources are listed, with reasons, in Supplementary Table S2.

Sixteen volume-equated trials, comprising 380 participants, contributed to the meta-analysis. Participant totals were 112 for the one-versus-two contrast, 106 for one-versus-three (counting the 12 participants of the within-subject trial, Neves et al., 2022, which contributed 24 limbs), and 162 for two-versus-three. Five trials compared one versus two sessions per muscle per week, six compared one versus three, and five compared two versus three.

Samples spanned untrained and trained men, untrained older and postmenopausal women, and three mixed-sex cohorts (McLester et al., 2000; Thomas & Burns, 2016; Candow & Burke, 2007, in which 23 of 29 participants were women). Durations ranged from six to twelve weeks. Hypertrophy was assessed by ultrasound muscle thickness in ten trials, magnetic resonance imaging cross-sectional area in one, dual-energy x-ray absorptiometry lean mass in three, and skinfold-derived lean mass in two. Study characteristics are summarized in Table 1.

**Table 1.**
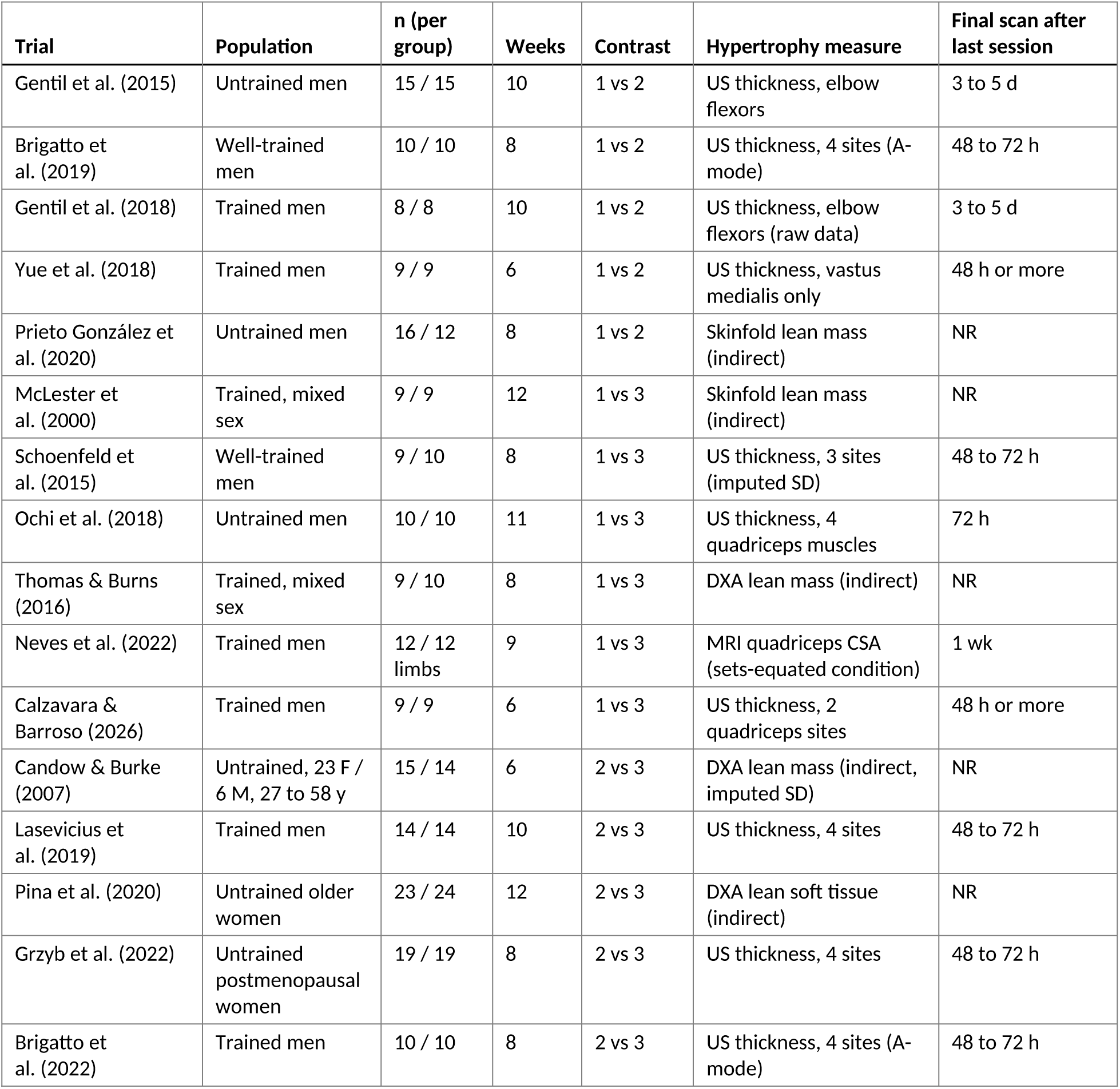
Characteristics of the included volume-equated frequency trials. Frequencies are expressed per muscle group per week. Gentil et al. (2018) presents muscle thickness only as an individual-values figure; its effect size was computed from the participant-level data published with the trial. Yue et al. (2018) contributes the vastus medialis site only, the one measured muscle trained once versus twice weekly (*Sections 2.2 and 2.4*). In Calzavara and Barroso (2026), weekly sets and relative load were equated, but the three-session arm completed more repetitions to failure at the same load, so its achieved volume load was higher (by 58% in the back squat and 76% in the leg press). US, ultrasound; CSA, cross-sectional area; DXA, dual-energy x-ray absorptiometry; NR, not reported. In Yue et al. (2018) and Calzavara and Barroso (2026), imaging was at least 48 h aker the last session. In Brigatto et al. (2019, 2022) and Schoenfeld et al. (2015) the 48 to 72 h interval is stated for baseline imaging and applied to post-training imaging by the same sentence.

| **Trial** | **Population** | **n (per group)** | **Weeks** | **Contrast** | **Hypertrophy measure** | **Final scan after last session** |
|---|---|---|---|---|---|---|
| Gentil et al. (2015) | Untrained men | 15 / 15 | 10 | 1 vs 2 | US thickness, elbow flexors | 3 to 5 d |
| Brigatto et al. (2019) | Well-trained men | 10 / 10 | 8 | 1 vs 2 | US thickness, 4 sites (A-mode) | 48 to 72 h |
| Gentil et al. (2018) | Trained men | 8 / 8 | 10 | 1 vs 2 | US thickness, elbow flexors (raw data) | 3 to 5 d |
| Yue et al. (2018) | Trained men | 9 / 9 | 6 | 1 vs 2 | US thickness, vastus medialis only | 48 h or more |
| Prieto González et al. (2020) | Untrained men | 16 / 12 | 8 | 1 vs 2 | Skinfold lean mass (indirect) | NR |
| McLester et al. (2000) | Trained, mixed sex | 9 / 9 | 12 | 1 vs 3 | Skinfold lean mass (indirect) | NR |
| Schoenfeld et al. (2015) | Well-trained men | 9 / 10 | 8 | 1 vs 3 | US thickness, 3 sites (imputed SD) | 48 to 72 h |
| Ochi et al. (2018) | Untrained men | 10 / 10 | 11 | 1 vs 3 | US thickness, 4 quadriceps muscles | 72 h |
| Thomas & Burns (2016) | Trained, mixed sex | 9 / 10 | 8 | 1 vs 3 | DXA lean mass (indirect) | NR |
| Neves et al. (2022) | Trained men | 12 / 12 limbs | 9 | 1 vs 3 | MRI quadriceps CSA (sets-equated condition) | 1 wk |
| Calzavara & Barroso (2026) | Trained men | 9 / 9 | 6 | 1 vs 3 | US thickness, 2 quadriceps sites | 48 h or more |
| Candow & Burke (2007) | Untrained, 23 F / 6 M, 27 to 58 y | 15 / 14 | 6 | 2 vs 3 | DXA lean mass (indirect, imputed SD) | NR |
| Lasevicius et al. (2019) | Trained men | 14 / 14 | 10 | 2 vs 3 | US thickness, 4 sites | 48 to 72 h |
| Pina et al. (2020) | Untrained older women | 23 / 24 | 12 | 2 vs 3 | DXA lean soft tissue (indirect) | NR |
| Grzyb et al. (2022) | Untrained postmenopausal women | 19 / 19 | 8 | 2 vs 3 | US thickness, 4 sites | 48 to 72 h |
| Brigatto et al. (2022) | Trained men | 10 / 10 | 8 | 2 vs 3 | US thickness, 4 sites (A-mode) | 48 to 72 h |

Significant within-group hypertrophy was reported in at least one arm in fourteen of the sixteen trials; the two indirect-measure one-versus-three trials (McLester et al., 2000; Thomas & Burns, 2016) reported no significant within-group change in either arm, Lasevicius et al. (2019) reported growth in the elbow extensors and quadriceps but not the elbow flexors, and in Gentil et al. (2018) and Prieto González et al. (2020) only the once-weekly arm changed significantly.

Independent re-screening and re-extraction of seven trials of the original set, purposively extended to cover the two trials flagged at screening in the original set for absent dispersion data (Gentil et al., 2018; Schoenfeld et al., 2015) and, among the randomly selected trials, one of the trials rated at high overall risk of bias (McLester et al., 2000), confirmed all seven eligibility decisions and all 35 extracted categorical fields (100% agreement throughout; Cohen’s kappa undefined under zero disagreement).

For the four trials added in October 2026, the second reviewer agreed with every coded design item and every extracted value checked against the trial reports; the participant-level changes for Gentil et al. (2018) were computed from the trial’s published data file and were not recomputed by the second reviewer. His notes led to two clarifications: in Prieto González et al. (2020) the split arm trained the upper body on one day and the trunk and lower body on the other (stated correctly in the Spanish text, whereas the English text and table headings are inconsistent), and in Calzavara and Barroso (2026) weekly sets were equated but achieved volume load was not (Table 1).

Independent recomputation of the change-score Hedges g for one trial (Gentil et al., 2015) reproduced the effect size to two decimal places (g = −0.20), and independent verification of a reported attrition statistic for a second trial (McLester et al., 2000) reproduced the figure exactly (28%). The check verified screening, extraction and derived statistics rather than re-appraising risk of bias, so it is a data-accuracy verification rather than a full dual-independent review. The recomputation of all sixteen pooled effect sizes from the trial reports, in code separate from the authors’ (Section 2.3), reproduced every value reported here.

### 3.2. Pooled effect of frequency on hypertrophy

Across the 16 trials in the pooled analysis, the point estimate for the effect of frequency on hypertrophy was trivial and non-significant (g = −0.06; 95% CI −0.24 to 0.13; p = 0.54). The interval includes zero; its upper bound lies within the trivial bound of |g| = 0.2 and its lower bound just crosses it, so a small effect favoring the higher frequency is not excluded, whereas a small effect favoring the lower frequency is. The truncated HKSJ interval was slightly wider (−0.26 to 0.14). The trials agreed more closely than chance alone would produce (Q = 8.45 on 15 degrees of freedom, p = 0.90; I² = 0%; τ² = 0), and the prediction interval (−0.26 to 0.14) was therefore close to the confidence interval. With 16 small trials and wide within-trial confidence intervals, I² has low power to detect heterogeneity and the between-trial variance is prone to a boundary estimate of zero, so the I² = 0% indicates that any between-trial variability is small relative to the within-trial confidence intervals rather than that all trials share an identical true effect, and the prediction interval should be read with the same reservation.

Contrast-specific estimates were consistent with the overall finding: one versus two sessions, g = +0.07 (95% CI −0.29 to 0.43; five trials, 112 participants); one versus three, g = −0.23 (95% CI −0.57 to 0.10; six trials, 106 participants); and two versus three, g = −0.01 (95% CI −0.29 to 0.27; five trials, 162 participants). None reached significance; the one-versus-two and two-versus-three estimates were trivial and the one-versus-three estimate small, and the contrast-specific intervals are wide. Seven trials returned point estimates beyond the trivial bound, two favoring the lower frequency (Gentil et al., 2018, +0.76; Lasevicius et al., 2019, +0.42) and five the higher (Neves et al., 2022, −0.62, the within-subject trial; McLester et al., 2000, −0.33, an indirect measure; Schoenfeld et al., 2015, −0.32, with imputed dispersion; Brigatto et al., 2022, −0.26; Grzyb et al., 2022, −0.21), all with intervals that include zero. Because the overall estimate pools contrasts that differ in frequency separation (a one-versus-three step spans two increments, a two-versus-three step one), the contrast-specific estimates are the more directly interpretable quantities, and the overall g is reported as a summary. Per-trial effects and the pooled diamonds are displayed in Figure 2, and the per-trial estimates are listed in Table 2.

**Figure 2.**
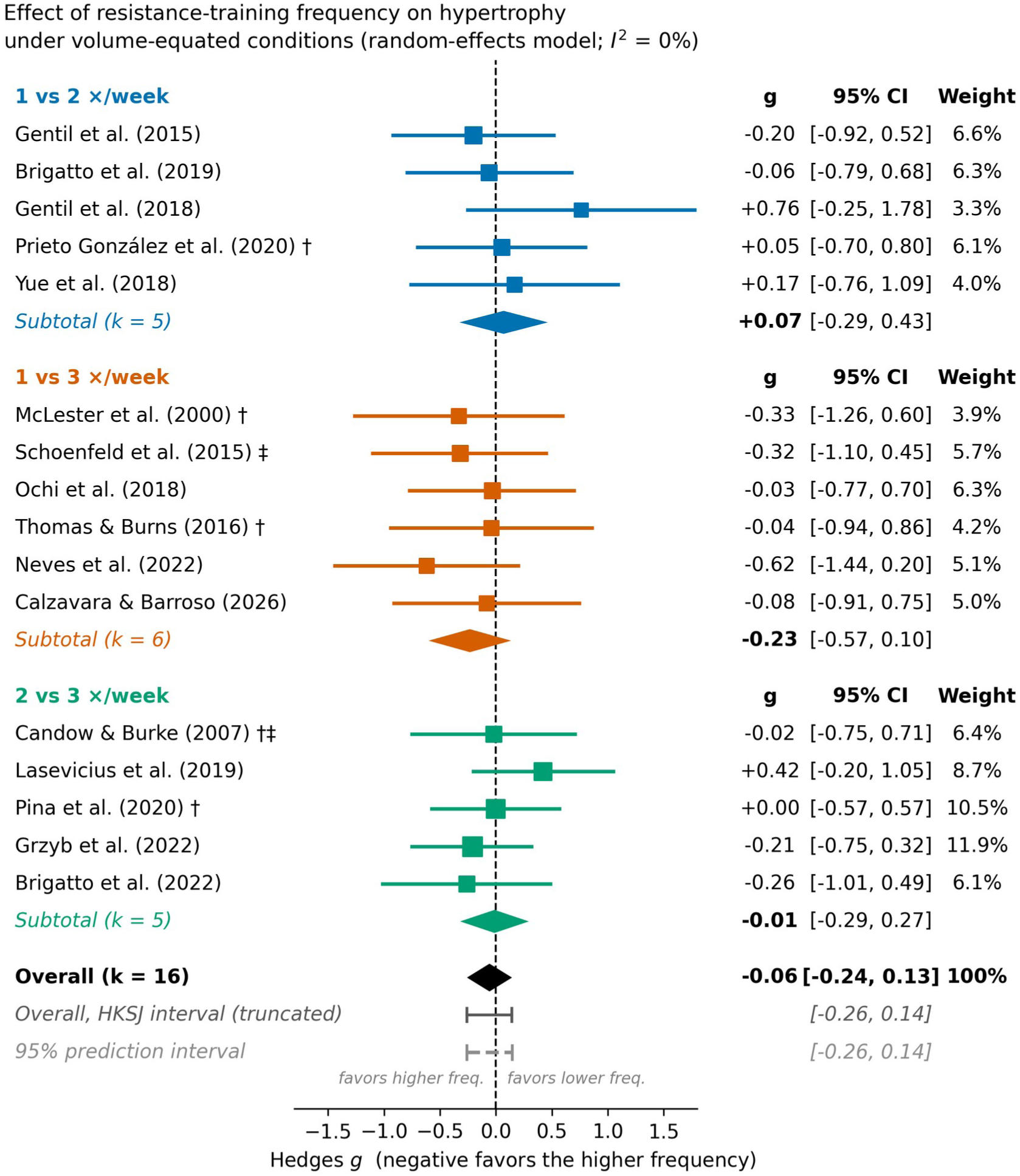
Forest plot of the change-score standardized mean difference (Hedges g) for hypertrophy under volume-equated conditions, grouped by frequency contrast. Negative values favor the higher frequency. Squares are per-trial estimates sized by inverse-variance weight; diamonds are random-effects pooled estimates by contrast and overall, with Wald 95% confidence intervals. The dashed vertical line marks no difference between frequencies (g = 0). The truncated HKSJ interval and the 95% prediction interval for the overall estimate are shown below the overall diamond. † indirect measure (skinfold or DXA lean mass); ‡ imputed change-score variance. The three contrast-specific subgroup diamonds are shown within this plot; a side-by-side paneled version is provided in the Supplementary Material (Figure S2).

**Table 2.**
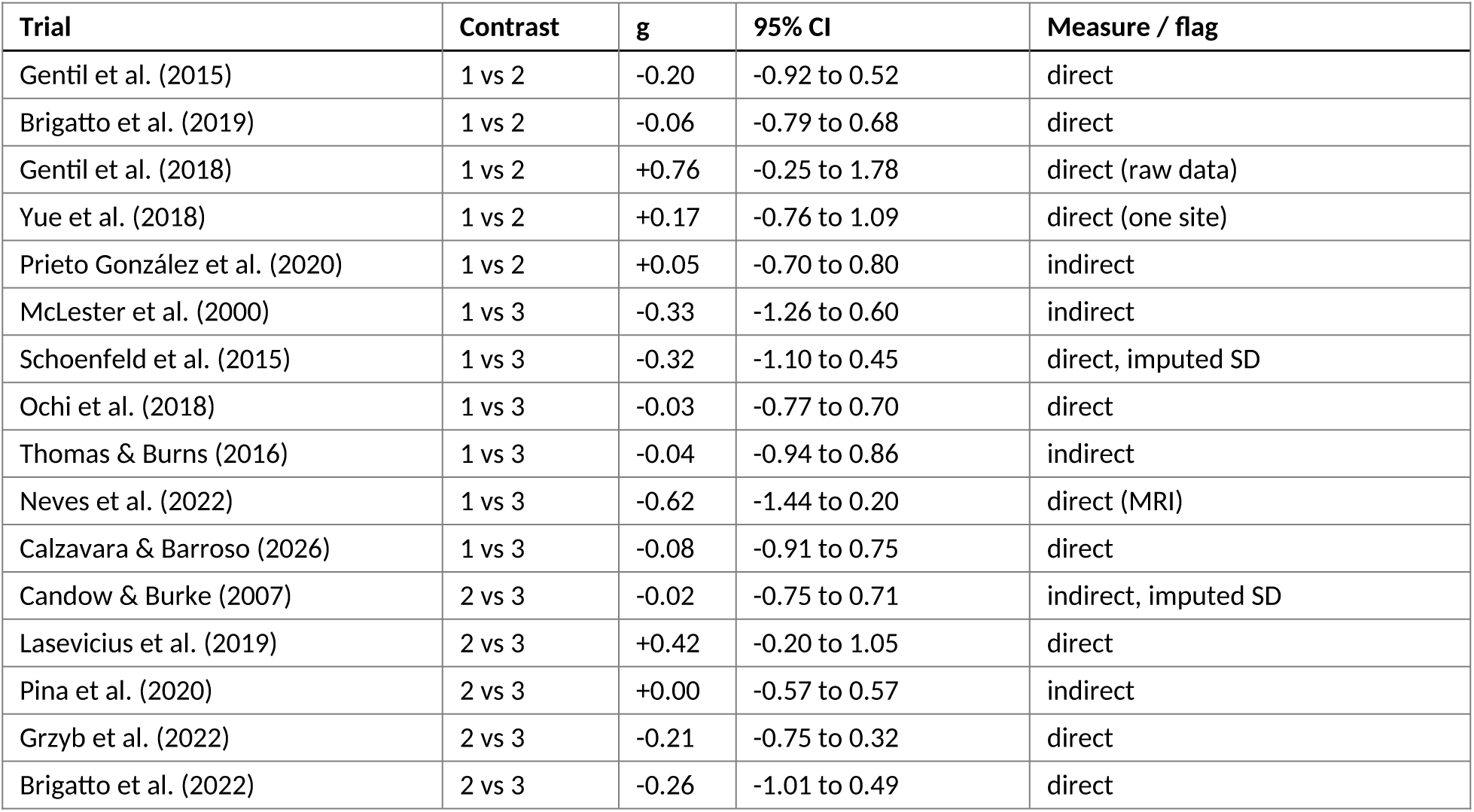
Per-trial aggregated effect sizes (Hedges g, change-score standardized mean difference; negative favors higher frequency).

### 3.3. Sensitivity analyses

The pooled point estimate remained trivial under every sensitivity analysis of the full set and moved by no more than 0.05. Restricting to direct measures of muscle size (eleven trials) returned g = −0.07 (95% CI −0.29 to 0.16); removing the two trials with imputed variance (fourteen trials) returned g = −0.04 (95% CI −0.24 to 0.15); and excluding the three trials rated at high risk of bias overall (McLester et al., 2000; Thomas & Burns, 2016; Prieto González et al., 2020) returned g = −0.05 (95% CI −0.25 to 0.15). The four direct-measure one-versus-three trials (Schoenfeld et al., 2015, −0.32; Ochi et al., 2018, −0.03; Neves et al., 2022, −0.62; Calzavara & Barroso, 2026, −0.08) pooled to g = −0.25 (95% CI −0.65 to 0.14).

Using the volume-load-equated condition of Neves et al. (2022) instead of its sets-equated condition (Section 2.5), as the first version of this review did, gives that trial g = +0.04 (95% CI −0.76 to 0.84) and returns g = −0.02 (95% CI −0.21 to 0.16) overall, −0.12 (95% CI −0.45 to 0.21) for one versus three sessions and −0.02 (95% CI −0.24 to 0.21) for the direct-measure trials; omitting the trial altogether returns g = −0.03 (95% CI −0.22 to 0.16) overall and −0.15 (95% CI −0.52 to 0.21) for one versus three. The choice of condition therefore moves the one-versus-three estimate by about 0.1 and the overall estimate by less than 0.04, in each case within the trivial range.

Trained cohorts (ten trials) returned g = −0.04 (95% CI −0.29 to 0.22) and untrained cohorts (six trials) g = −0.08 (95% CI −0.35 to 0.19); the difference was +0.04 (95% CI −0.33 to 0.41; p = 0.82). Removing the four trials added in the October 2026 update returned the corrected estimate for the original twelve trials (g = −0.11, 95% CI −0.31 to 0.10), and removing Gentil et al. (2018) alone, the trial most favoring the lower frequency, returned g = −0.09 (95% CI −0.27 to 0.10).

All eleven trials using direct measures of muscle size took the final measurement 48 hours or more after the last training session (48 to 72 hours in five, at least 48 hours in Yue et al. (2018) and Calzavara and Barroso (2026), 72 hours in Ochi et al. (2018), three to five days in Gentil et al. (2015, 2018) and one week in Neves et al. (2022)); seven of them state that the interval was chosen to allow training-induced swelling to resolve. None of the five trials using indirect measures reported the interval. Restricting the analysis to the eleven trials with a documented interval of 48 hours or more therefore coincides with the direct-measure analysis above (g = −0.07, 95% CI −0.29 to 0.16; I² = 0%), and the five trials without a documented interval gave g = −0.04 (95% CI −0.37 to 0.29). No trial reported echo intensity; Ochi et al. (2018) measured muscle stiffness by shear-wave elastography 72 hours after the last session as an index of muscle damage and found no change in either group and no difference between groups.

The pre-post correlation is the denominator of every effect size derived from pre and post standard deviations (nine of the sixteen trials), so it changes the per-trial estimates rather than canceling: at r = 0.5 these estimates shrink by a factor of 0.77 and at r = 0.9 they grow by 1.73, so that at r = 0.9 McLester et al. (2000) becomes −0.57 and Lasevicius et al. (2019) +0.73. Because the affected trials lie on both sides of zero, the pooled estimate moved little (r = 0.5, g = −0.05, 95% CI −0.24 to 0.13, Q = 7.21; r = 0.9, g = −0.07, 95% CI −0.26 to 0.12, Q = 14.48 on 15 degrees of freedom), but the apparent agreement among trials at r = 0.7 is partly a consequence of the assumed correlation.

For whole-body lean mass the test-retest correlation is probably higher than 0.9; applying r = 0.95 and 0.98 to the three indirect trials whose effects depend on it (McLester et al., 2000; Candow & Burke, 2007; Prieto González et al., 2020) returned g = −0.07 (95% CI −0.26 to 0.11) and g = −0.08 (95% CI −0.27 to 0.10), with McLester et al. (2000) at −0.81 and −1.28, an implausibly large effect that reflects the assumption rather than the data.

Varying the within-trial correlation of the multi-site composites between 0.3 and 1.0 left the pooled estimate at −0.06, with the interval widening as the correlation rose (95% CI −0.23 to 0.11 at 0.3; −0.26 to 0.14 at 1.0), because a higher correlation reduces the weight of the multi-site trials. Adding Tavares et al. (2017) (g = −0.21, 95% CI −1.05 to 0.62, from the trial’s reported change-score intervals) returned g = −0.07 (95% CI −0.25 to 0.12) overall and g = +0.03 (95% CI −0.31 to 0.36) for the six one-versus-two trials.

Leave-one-out analysis ranged from −0.03 (omitting Neves et al., 2022) to −0.10 (omitting Lasevicius et al., 2019; 95% CI −0.30 to 0.09), with no omission moving the estimate beyond the trivial bound (Figure 3). Unlike the twelve-trial analysis, no omission brought even the unmodified HKSJ interval to exclude zero (with Lasevicius et al. omitted, −0.24 to 0.04). Taken together, the sensitivity analyses of the full set show a pooled point estimate that is trivial under every specification examined, with intervals that admit a small effect favoring the higher frequency; in the smaller subsets the point estimates reach the small-effect bound on that side (the four direct-measure one-versus-three trials, −0.25; the four blinded direct-measure trials, −0.20, Section 3.7) and the intervals admit small effects in either direction.

**Figure 3.**
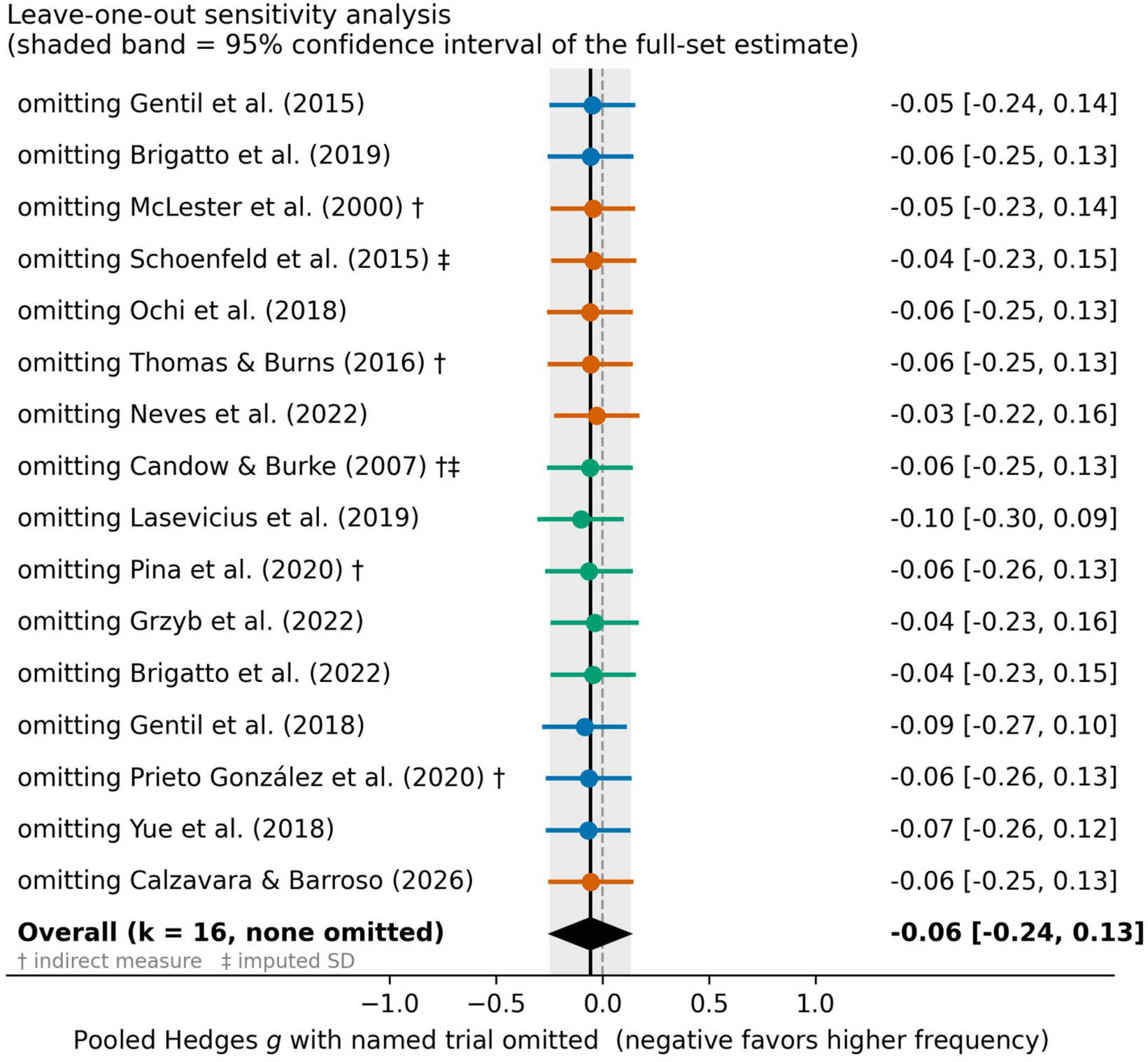
Leave-one-out sensitivity analysis. Each row shows the pooled estimate (Wald 95% CI) when the named trial is omitted; the shaded band is the 95% CI of the full-set estimate. The pooled estimate remains trivial across all omissions (range −0.03 to −0.10); Lasevicius et *al. (2019), Gentil et al. (2018) and Neves et al. (2022) are the most influential omissions*.

### 3.4. Moderation by intervention duration

Because intervention length varied from six to twelve weeks, a random-effects meta-regression tested whether duration moderated the frequency effect. The slope was small and non-significant (+0.02 g per week, 95% CI −0.08 to 0.12; p = 0.75), and residual heterogeneity remained at zero. The fitted model implied a predicted g of approximately −0.10 at six weeks and −0.01 at twelve weeks. The regression could, however, have detected only a slope of about 0.14 g per week with 80% power, a change of about 0.8 g across the six-to-twelve-week range, so it provides no evidence either way about smaller duration effects; with a narrow span of durations and no heterogeneity to explain, it is best read as finding no duration effect large enough to be detected within the common six-to-twelve-week window.

### 3.5. Moderation by training volume

Two exploratory random-effects meta-regressions tested whether training volume moderated the frequency effect. The first used the per-session set volume for the measured muscle in the lower-frequency arm, the arm worked hardest in each session (range three to eighteen sets); the second used the weekly set volume for the measured muscle (range three to thirty-six sets). Sets were counted with one rule, as every set of an exercise in which the measured muscle is a prime mover, so that the eighteen-set quadriceps session of Lasevicius et al. (2019), the sixteen-set sessions of Brigatto et al. (2019) and the twelve-set elbow-flexor sessions of Gentil et al. (2015, 2018) are counted on the same basis (Supplementary Table S4).

Neither moderated the frequency effect. The per-session slope was small and non-significant (+0.024 g per set, 95% CI −0.016 to 0.063; p = 0.24), as was the weekly slope (+0.016 g per set, 95% CI −0.006 to 0.038; p = 0.15), and residual heterogeneity remained at zero in both models. Both slopes are positive, meaning that the trials with larger sessions leaned, if anything, toward the lower frequency, which is the opposite of the direction a per-session ceiling would produce; but the regressions could have detected only slopes of about 0.06 g per set (per-session) and 0.03 g per set (weekly) with 80% power, each a change of about 0.9 to 1.0 g across the observed range, so they provide no evidence either way about smaller volume effects.

Three trials tied for the lowest per-session volume (three sets); the nine trials at nine or more sets per session included the three that most favored the higher frequency (Neves et al., 2022, −0.62; Schoenfeld et al., 2015, −0.32; Brigatto et al., 2022, −0.26) as well as the two that most favored the lower frequency (Gentil et al., 2018, +0.76; Lasevicius et al., 2019, +0.42) (Figure 4). Per-session and weekly volume were themselves correlated (r = 0.81). These analyses also cannot separate the influence of training volume from that of proximity to failure, which was prescribed inconsistently across the included trials (Supplementary Table S1).

**Figure 4.**
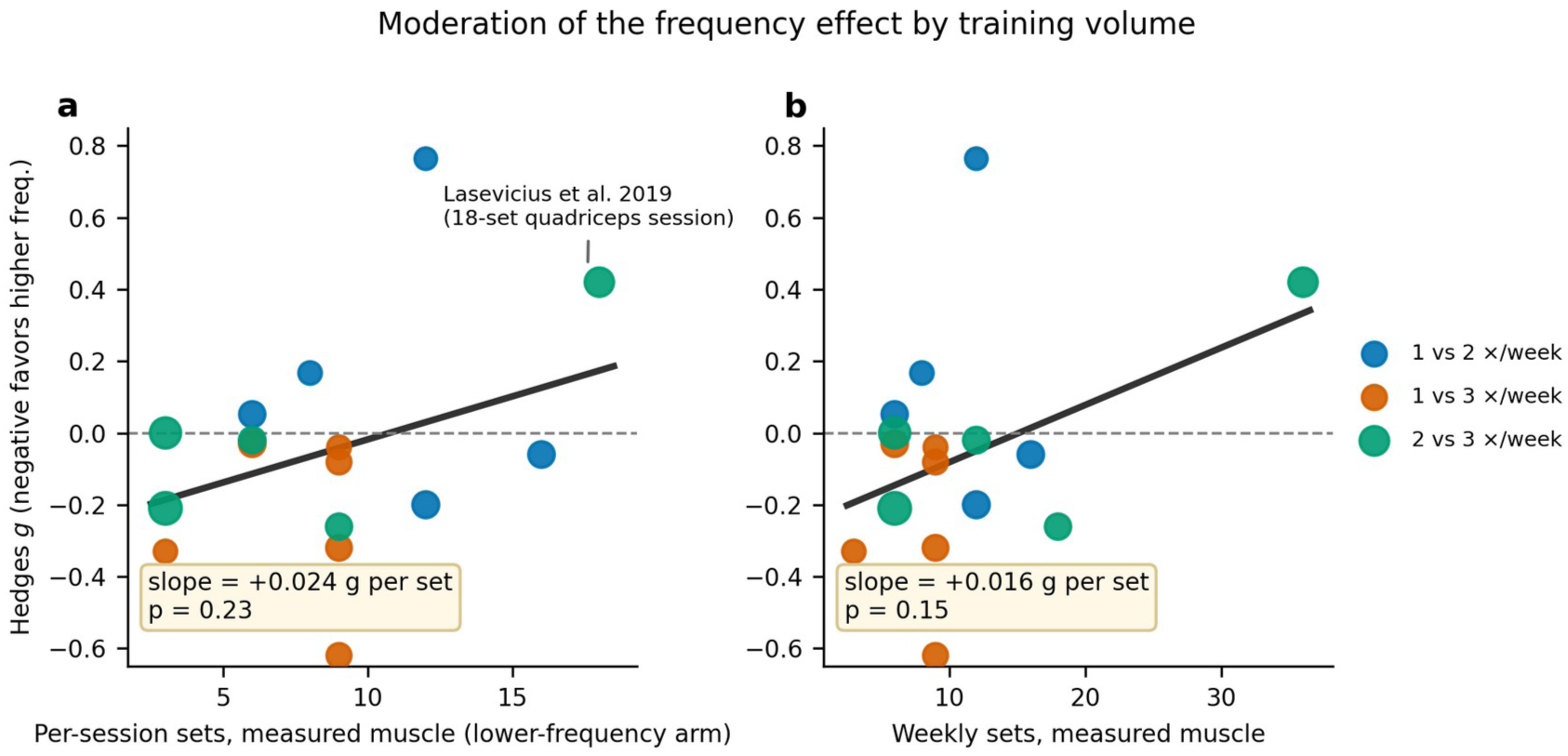
Frequency effect (Hedges g) regressed on training volume across the sixteen pooled trials, with sets counted for the measured muscle under the rule in Supplementary Table S4. (a) Per-session set volume of the lower-frequency arm (slope +0.024 g per set, 95% CI −0.016 to 0.063, p = 0.24); the highest per-session point (18 sets) is the quadriceps session of the two-session arm of Lasevicius et al. (2019). (b) Weekly set volume for the measured muscle (slope *+0.016 g per set, 95% CI −0.006 to 0.038, p = 0.15). Residual heterogeneity is zero in both models; the detectable slope at 80% power is about 0.9 to 1.0 g across the observed range in each panel. Negative values favor the higher frequency; point size reflects inverse-variance weight*.

Repetition range and relative load were examined as a further exploratory moderator. The included trials clustered in the moderate range: ten of the sixteen prescribed eight to twelve repetitions and two more were close (six to twelve in Prieto González et al., 2020; sets to failure at 80% of one-repetition maximum in Calzavara & Barroso, 2026), with McLester et al. (2000) at the heavy, low-repetition end (a periodized three to ten range, mean approximately 6.5), Grzyb et al. (2022) at the light, high-repetition end (twenty to thirty repetitions), and Ochi et al. (2018) and Pina et al. (2020) at twelve to fifteen repetitions. Coding each trial by its mean prescribed repetitions, the meta-regression slope was −0.009 g per repetition (95% CI −0.046 to 0.028, p = 0.64), with residual heterogeneity at zero (Figure 5a); coding instead by approximate relative load gave the same result, a small non-significant slope (+0.004 g per percent of one-repetition maximum, 95% CI −0.026 to 0.033, p = 0.81; Figure 5b).

**Figure 5.**
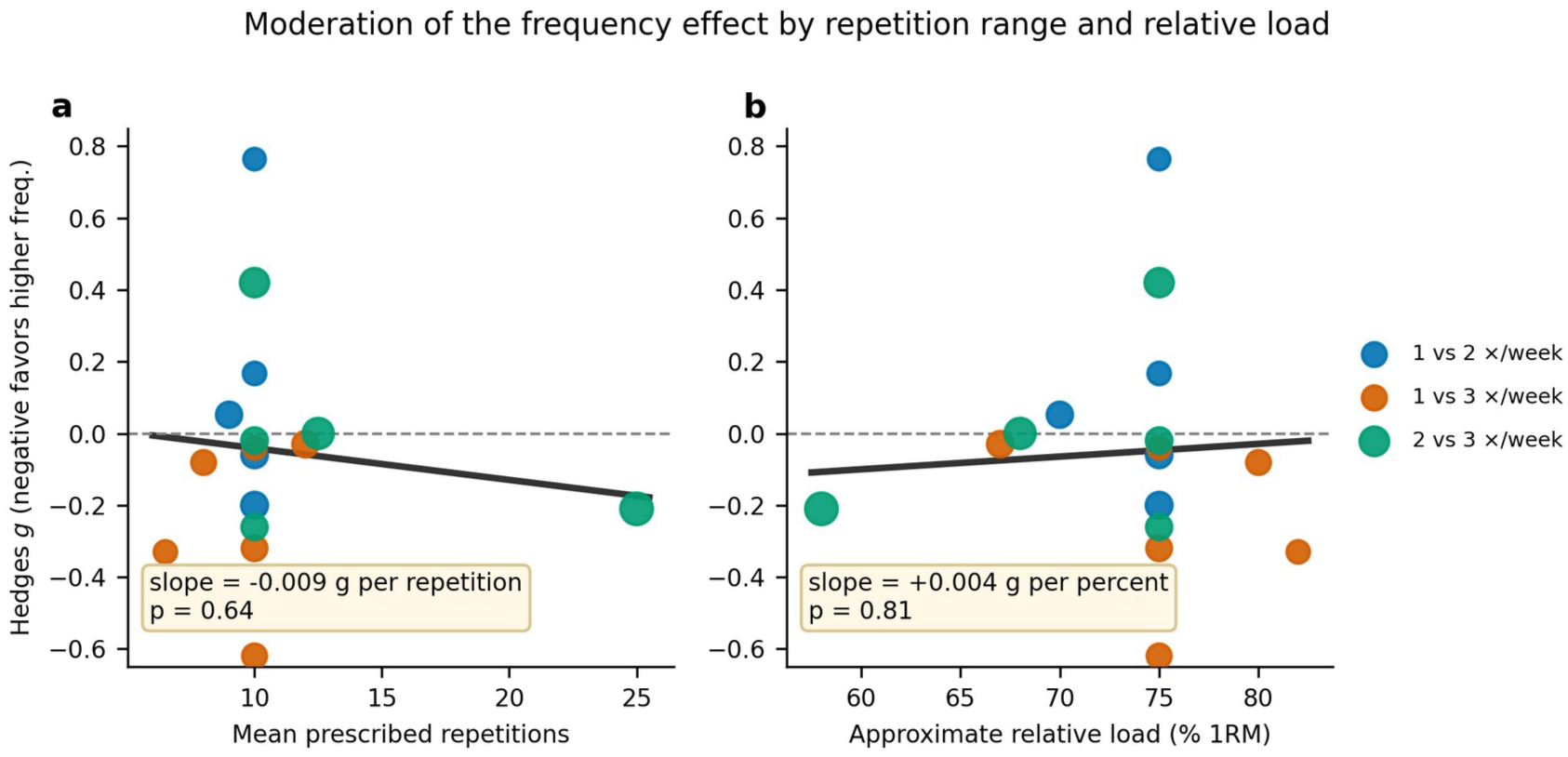
Frequency effect (Hedges g) regressed on loading characteristics across the sixteen pooled trials. (a) Mean prescribed repetitions (slope −0.009 g per repetition, 95% CI −0.046 to 0.028, p = 0.64). (b) Approximate relative load as a percentage of one-repetition maximum (slope +0.004 g per percent, 95% CI −0.026 to 0.033, p = 0.81). Both associations are small and non-significant, with residual heterogeneity at zero; the detectable slope at 80% power is about *1.0 g across the observed range in each panel. Negative values favor the higher frequency; point size reflects inverse-variance weight*.

The repetition-coded point estimate reversed sign when the single high-repetition trial was removed (slope +0.030, p = 0.68) and fell to near zero when the two extreme trials were removed together (slope +0.006, p = 0.95). The detectable slopes were about 0.053 g per repetition and 0.042 g per percent, each a change of about 1.0 g across the observed range. Repetition range and load therefore showed no moderating relationship with the frequency effect that these analyses could have detected; the narrow clustering of loads leaves them unable to detect anything but a large one.

### 3.6. Risk of bias

Risk of bias is summarized in Table 3. Most trials raised some concerns, chiefly from incomplete reporting of allocation concealment and the absence of prospective registration, and from unblinded outcome assessment. Three trials were judged at high risk overall. McLester et al. (2000) combined a 28% attrition with a per-protocol analysis and used a skinfold-derived estimate of lean mass, an indirect index of limited validity for hypertrophy, and Prieto González et al. (2020) likewise excluded 11 of 39 participants for non-adherence and analyzed per protocol, with a skinfold-derived lean-mass outcome and no blinding statement. Thomas and Burns (2016) allocated participants to conditions by demographic balancing rather than randomization; the high rating rests on allocation.

**Table 3.** Risk-of-bias appraisal (Cochrane RoB 2). L, low risk; S, some concerns; H, high risk. D1, randomization process; D2, deviations from intended interventions; D3, missing outcome data; D4, measurement of the outcome; D5, selection of the reported result. Gentil et al. (2015) and Gentil et al. (2018) each excluded four participants post hoc for non-compliance and are rated alike on D3; Calzavara and Barroso (2026) analyzed the 18 of 23 randomized participants who completed every session. Per-protocol analysis was rated high risk on D3 where more than a quarter of randomized participants were excluded (McLester et al., 2000; Prieto González et al., 2020) and some concerns otherwise. Neves et al. (2022) allocated limbs by counterbalancing without a concealment statement (D1). Candow and Burke (2007) and Yue et al. (2018) state blinded assessment (D4); Grzyb et al. (2022) make no blinding statement (D4). No trial reports prospective registration (D5).

| Trial | D1 | D2 | D3 | D4 | D5 | Overall |
|---|---|---|---|---|---|---|
| Gentil et al. (2015) | S | L | S | S | S | S |
| Brigatto et al. (2019) | S | L | L | S | S | S |
| Gentil et al. (2018) | S | L | S | S | S | S |
| Yue et al. (2018) | S | L | L | L | S | S |
| Prieto González et al. (2020) | S | S | H | H | S | H |
| McLester et al. (2000) | S | S | H | H | S | H |
| Schoenfeld et al. (2015) | S | L | L | S | S | S |
| Ochi et al. (2018) | S | L | L | L | S | S |
| Thomas & Burns (2016) | H | S | L | S | S | H |
| Neves et al. (2022) | S | S | L | L | S | S |
| Calzavara & Barroso (2026) | S | L | S | S | S | S |
| Candow & Burke (2007) | S | L | L | L | S | S |
| Lasevicius et al. (2019) | S | L | S | S | S | S |
| Pina et al. (2020) | S | L | L | S | S | S |
| Grzyb et al. (2022) | S | L | L | S | S | S |
| Brigatto et al. (2022) | S | L | L | L | S | S |

Four trials report blinded assessment of a direct muscle-size measure: Ochi et al. (2018), Neves et al. (2022), Brigatto et al. (2022) and Yue et al. (2018); Grzyb et al. (2022) reports that one researcher performed all scans and another analyzed all images, without a blinding statement, Calzavara and Barroso (2026) state that the assessor was not blinded, and Candow and Burke (2007) describe a double-blind design for an indirect measure. The four blinded direct-measure trials pooled to g = −0.20 (95% CI −0.60 to 0.20).

In the one-versus-three contrast, the lean toward the higher frequency came mainly from Neves et al. (2022), rated at some concerns, with smaller contributions from McLester et al. (2000), rated at high risk, and Schoenfeld et al. (2015), rated at some concerns; removing the two high-risk trials moved the contrast estimate from −0.23 to −0.25. In the one-versus-two contrast, removing Prieto González et al. (2020) left the estimate at +0.07.

### 3.7. Certainty of evidence (GRADE)

The certainty of the pooled evidence was rated using the GRADE approach (Guyatt et al., 2008) for each of the three frequency contrasts. The full GRADE evidence profile is presented in Table 4. Randomized trials begin at high certainty; Thomas and Burns (2016), which allocated by demographic balancing, was treated as a randomized trial at high risk of bias rather than as non-randomized evidence. The evidence was downgraded for risk of bias (most trials raised some concerns; three were at high risk), for imprecision, and for indirectness.

**Table 4.** GRADE evidence profile: resistance training frequency and muscle hypertrophy by contrast type.

| Contrast | k (n) | Pooled g (95% CI) | RoB | Incons. | Indir. | Imprec. | Pub. bias | Certainty | Interpretation |
|---|---|---|---|---|---|---|---|---|---|
| 1 vs 2 | 5 (112) | +0.07 (-0.29 to 0.43) | -1 | 0 | -1 | -1 | none | VERY LOW | Interval spans small effects in either direction; one high-risk skinfold trial carries 23% of the weight |
| 1 vs 3 | 6 (106) | -0.23 (-0.57 to 0.10) | -1 | 0 | -1 | -1 | none | VERY LOW | Small lean toward higher frequency, chiefly from the within-subject MRI trial; direct-measure subset -0.25 (-0.65 to 0.14) |
| 2 vs 3 | 5 (162) | -0.01 (-0.29 to 0.27) | -1 | 0 | 0 | -1 | none | LOW | Most precise contrast; trivial point estimate, small effects either way not excluded |
| Overall | 16 (380) | -0.06 (-0.24 to 0.13) | -1 | 0 | 0 | -1 | none | LOW | Trivial point estimate; the interval crosses the small-effect bound only on the side favoring the higher frequency |

Abbreviations: k, number of trials; n, total participants; CI, confidence interval (Wald); RoB, risk of bias; Incons., inconsistency; Indir., indirectness; Imprec., imprecision; Pub. bias, publication bias. Downgrades: 0, not serious; -1, serious; -2, very serious; none, undetected. GRADE certainty ratings: High (no downgrade), Moderate (-1), Low (-2), Very Low (-3 or more). Optimal information size for g = 0.2 at 80% power: about 785 participants. The weight-based reasoning for the risk-of-bias and indirectness downgrades is given in Section 3.7.

The optimal information size for the smallest effect size of interest (g = 0.2, 80% power, two-sided alpha 0.05) is about 785 participants; the pool contains 380, and the contrasts 112, 106 and 162. Imprecision was rated serious for every contrast and for the overall estimate: each interval includes a small effect favoring the higher frequency and, except for the one-versus-three contrast and the overall estimate, a small effect favoring the lower frequency.

Risk of bias was rated serious throughout. The high-risk trials carry 23% of the one-versus-two weight (Prieto González et al., 2020), 27% of the one-versus-three weight (McLester et al., 2000; Thomas & Burns, 2016) and 14% of the overall weight, and removing them moved no estimate by more than about 0.02. Indirectness was rated serious where a skinfold-derived lean-mass estimate, a weaker surrogate for muscle-specific hypertrophy than the dual-energy x-ray absorptiometry measures of the other indirect trials (Candow & Burke, 2007; Pina et al., 2020; Thomas & Burns, 2016), carries a substantial share of a contrast’s weight: 23% of the one-versus-two weight (Prieto González et al., 2020) and 13% of the one-versus-three weight (McLester et al., 2000), against 10% of the overall weight (not downgraded); dual-energy x-ray absorptiometry alone was not treated as grounds for downgrading.

Inconsistency and publication bias were not grounds for downgrading (I² = 0%; Egger’s test non-significant). On balance, the GRADE profiles indicate low to very low certainty that frequency has no more than a trivial effect on hypertrophy at equated weekly volume within the one-to-three-session range; small effects are not excluded. The direction of the finding agrees with the overview of reviews by Currier et al. (2026), which found no influence of frequency on hypertrophy when total volume was equated.

### 3.8. Publication bias

A formal publication bias assessment was conducted using a funnel plot and Egger’s regression test (Egger et al., 1997). The funnel plot (Figure 6) and contour-enhanced funnel plot (Figure 7) are presented below. With k = 16 trials, formal tests have limited statistical power; the assessment is therefore exploratory. A further caveat applies to the regression test itself: because a standardized mean difference is mathematically correlated with its own standard error, Egger’s test on SMD data can show inflated type-I error and detect asymmetry that reflects this artifact rather than selective reporting, so a non-significant test here is reassuring but not decisive. The funnel also pools the three frequency contrasts, which differ in their expected effect, so apparent scatter partly reflects contrast membership rather than small-study effects.

**Figure 6.**
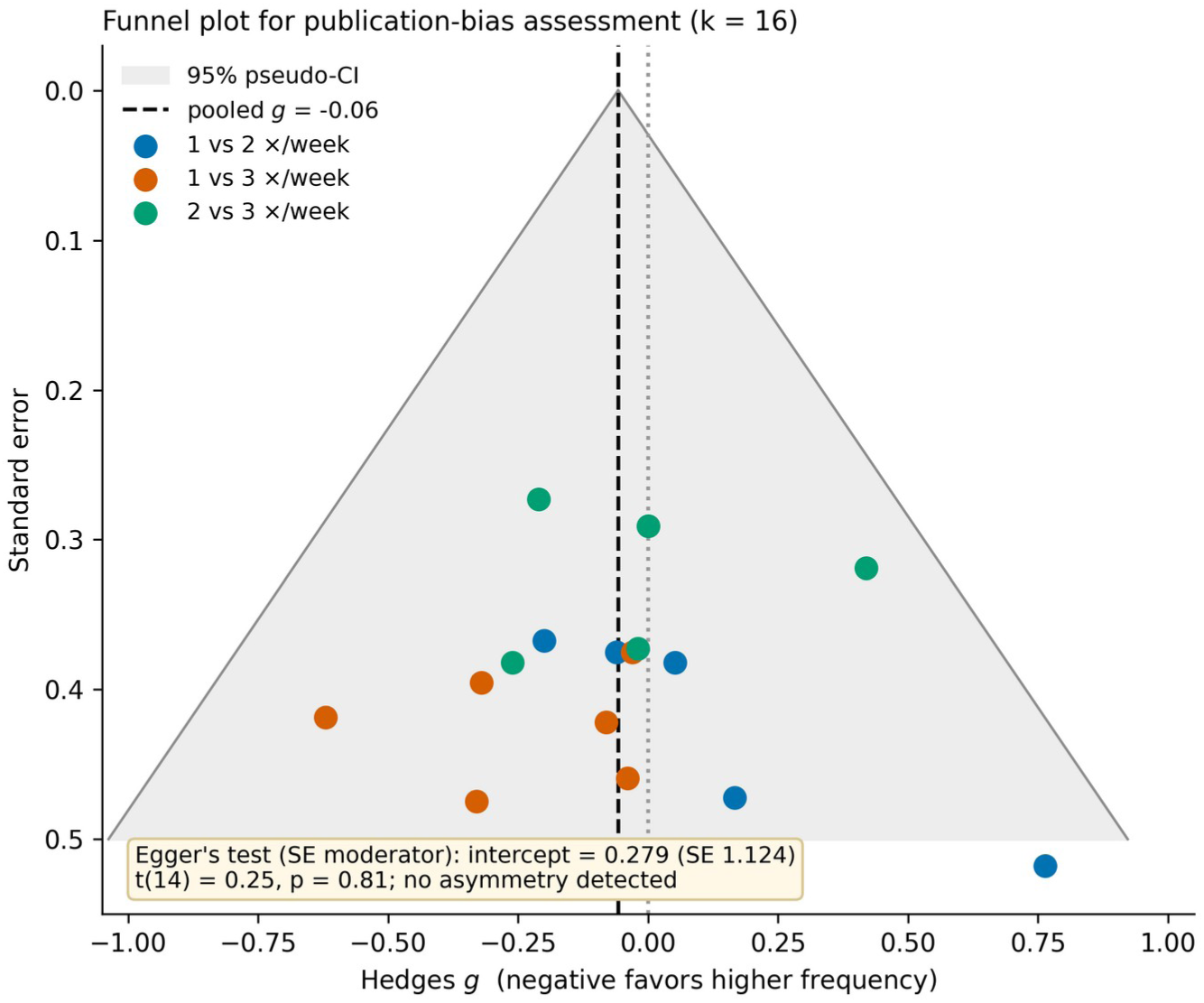
Funnel plot for publication-bias assessment (k = 16). Each point is one trial, colored by contrast. The dashed vertical line marks the pooled estimate (g = −0.06); the shaded region is the 95% pseudo-confidence funnel. The plot is approximately symmetric (Egger’s test with a standard-error moderator: intercept = +0.28, SE = 1.12, t(14) = 0.25, p = 0.81).

**Figure 7.**
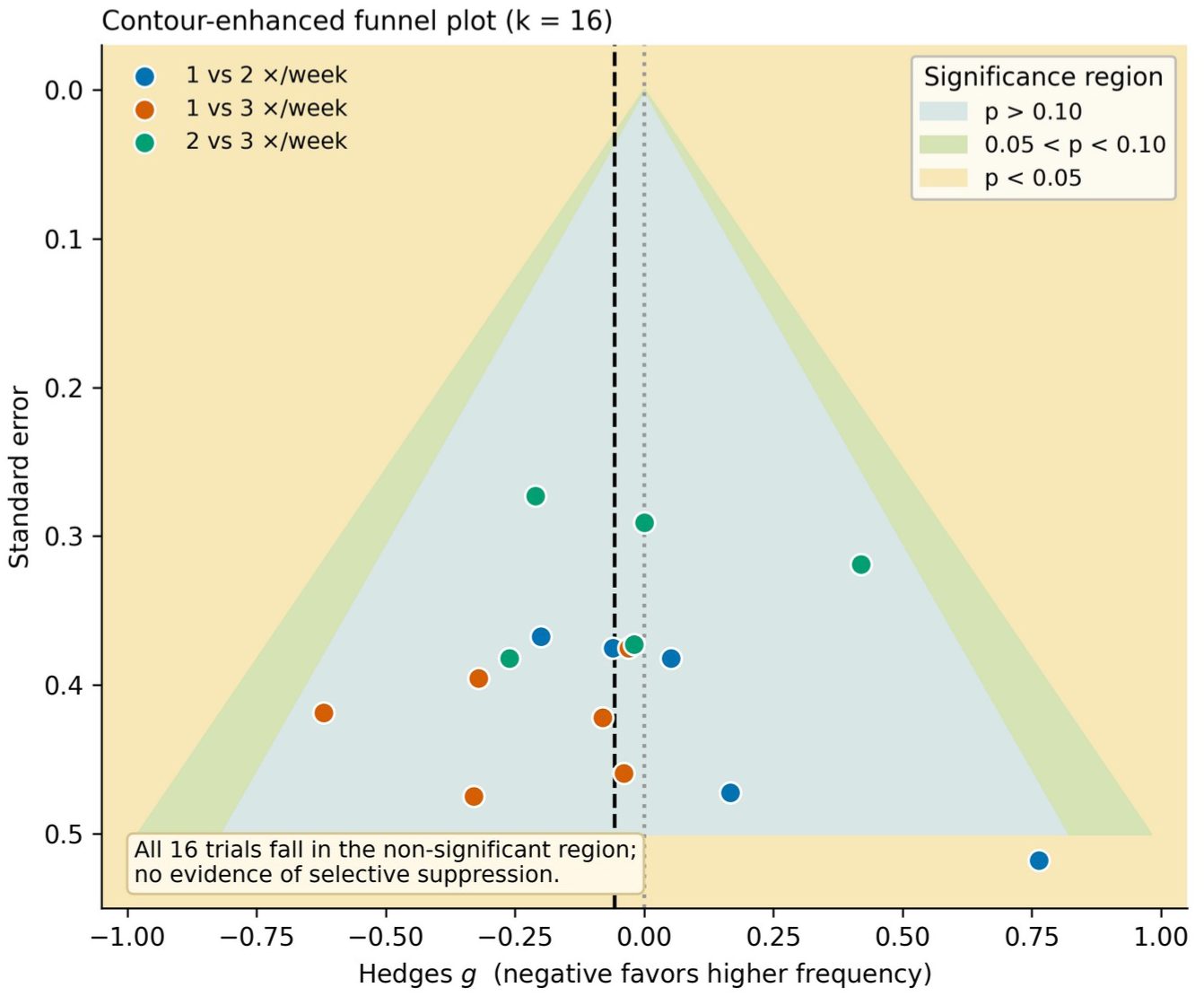
Contour-enhanced funnel plot (k = 16). Shaded regions indicate significance thresholds (blue: p > 0.10; green: 0.05 < p < 0.10; yellow: p < 0.05). All sixteen trials fall within the non-significant region.

Egger’s regression test yielded an intercept of +0.28 (SE = 1.12; t(14) = 0.25; p = 0.81) with the standard-error moderator and +0.44 (SE = 2.35; t(14) = 0.18; p = 0.86) with the sample-size-based precision term, providing no statistical evidence of funnel plot asymmetry under either specification. The positive intercepts indicate a slight tendency for smaller trials to show more positive (lower-frequency-favoring) effects, which is small and non-significant. All 16 trials fall within the non-significant region of the contour-enhanced funnel plot (Peters et al., 2008). While the limited number of trials means that formal tests cannot definitively exclude publication bias, the approximately symmetric funnel and non-significant Egger’s test are reassuring. Five of the sixteen trials were found outside the database searches, in reference lists or in the earlier search (Section 3.1), so trials of this design in less widely indexed journals are not reliably retrieved by database strings, and further unretrieved trials cannot be excluded.

## 4. Discussion

### 4.1. Main findings

Across 16 volume-equated resistance-training trials, our synthesis indicates that, within the ecologically common range of one to three sessions per muscle per week, altering frequency while holding weekly volume constant does not produce large or clearly detectable differences in hypertrophy. The pooled effect was close to zero, with a confidence interval that admits at most a small effect favoring the higher frequency and excludes a small effect favoring the lower. The point estimate was trivial in the ultrasound-thickness and lean-mass subsets and in trained and untrained cohorts (the single MRI trial, Neves et al., 2022, returned a moderate effect favoring the higher frequency, discussed in Section 4.4), although the older and postmenopausal cohorts are represented by two two-versus-three trials only. Read together, these findings are consistent with the view that weekly volume is the primary modifiable driver of hypertrophy (Pelland et al., 2026; Schoenfeld et al., 2017), and that frequency within the one-to-three-session range acts chiefly as a distribution variable.

### 4.2. Comparison with previous meta-analyses

Our conclusion agrees with the volume-equated syntheses that precede it. Schoenfeld et al. (2016) reported an advantage for training a muscle twice rather than once per week in seven volume-equated comparisons (0.19, 95% CI 0.11 to 0.28), mostly in untrained participants and with indirect measures; with 13 volume-equated studies and a different effect-size model, Schoenfeld et al. (2019) revised this to a trivial and non-significant difference (0.07, 95% CI −0.02 to 0.17), in agreement with the present pooled estimate. Grgic et al. (2018) found for strength that the frequency effect was no longer significant once volume was matched. Pelland et al. (2026), modeling the weekly set volume dose-response, reported a frequency coefficient for hypertrophy that was inconsistent across analyses and compatible with negligible effects, and their analysis restricted to the fifteen studies with direct within-study frequency comparisons did not indicate an effect of frequency; their strength model, by contrast, favored higher frequency, with practice of the strength test among the candidate explanations. The present review is a narrower, trial-level counterpart to those results: it is restricted to the one-to-three-session range, uses one composite effect per trial, and states what its analyses could and could not detect.

The overview of reviews by Currier et al. (2026) likewise concluded that resistance training frequency did not influence hypertrophy when total volume was equated. By focusing specifically on per-muscle, volume-equated hypertrophy trials in the one-to-three-session range, deriving one change-score effect size per trial directly from the trial reports, and integrating GRADE certainty ratings, the present review complements these broader syntheses with an estimate for the schedules that practice most often uses.

The most instructive comparison is with Tao et al. (2026), which reached a similar conclusion for muscle mass in volume-equated comparisons (equal-volume subgroup g = 0.01) despite using different analytic choices. Their approach to deriving change-score dispersion and coding frequency contrasts yielded substantially larger per-trial standardized mean differences and higher apparent heterogeneity than in the present analysis; these differences arise from extraction and contrast-coding choices rather than from conflicting underlying data, and a detailed cross-check is provided in the Supplementary Material (Section S13 and Table S7). Tao et al. also reported a secondary signal favoring higher frequency for upper-body muscle, and the largest subgroup estimate in Schoenfeld et al. (2019) was for the lower body (0.15, p = 0.08); because the present review averages sites within each trial, it cannot examine region, which is noted as a limitation (Section 4.6) and a design point for future trials (Section 4.7).

The present synthesis shares eight of its sixteen trials (Gentil et al., 2015, 2018; Brigatto et al., 2019; Schoenfeld et al., 2015; Ochi et al., 2018; Yue et al., 2018; Lasevicius et al., 2019; Neves et al., 2022) with the open corpus of direct hypertrophy trials analyzed in a companion model-based re-analysis by the present authors (Boutagy & McKean, 2026; posted as a preprint and under review) of the extracted dataset deposited with Pelland et al. (2026); that corpus codes the sets-equated condition of Neves et al. (2022), as the present analysis now does (Section 2.5). That analysis estimated the coefficient for partitioning a fixed weekly volume across sessions at −0.043 per unit ln frequency (95% CI −0.161 to 0.074) across the corpus and −0.063 (95% CI −0.194 to 0.069) within studies, and a difference of −0.005 SMCR (95% CI −0.165 to 0.155) in its fifteen within-study frequency contrasts at equal weekly volume, with the coefficient between −0.051 and −0.020 under five alternative specifications that address measurement timing and swelling; in that analysis negative values favor the lower frequency, the opposite of the sign convention used here. Agreement between the two papers is therefore partial triangulation rather than independent replication.

### 4.3. Frequency, volume and the hypertrophic response

A common objection to volume-equated frequency comparisons is that per-set hypertrophic returns diminish within a session, so distributing a fixed weekly volume across more sessions should increase the proportion of ‘high-value’ early sets and favor higher frequency. This reasoning draws on dose-response analyses of weekly volume (Schoenfeld et al., 2017; Pelland et al., 2026) and per-session set volume (Remmert et al., 2025), and could appear to predict a clear ranking, with three sessions ahead of two and both ahead of one at matched weekly sets. The present results do not show that ranking, and the trials in which it should be most visible, the nine whose lower-frequency arm performed nine to eighteen sets for the measured muscle in one session, show no frequency benefit (Figure 4).

Remmert et al. (2025), in a preprint, reported a gradually saturating per-session hypertrophy curve with a practical upper observed value of about 11 fractional sets per session, which they present as an observed range rather than an upper limit, while Pelland et al. (2026) found a frequency coefficient for hypertrophy compatible with negligible effects in a weekly volume model with no clear plateau. Taken together, these data are consistent with distributing moderate weekly volumes across one to three sessions changing the timing of the stimulus more than its magnitude, within the range studied and at the precision available, with the reservation that the present moderator analyses could detect only a large volume dependence (Section 3.5).

The geometric decay illustration in Supplementary Notes (Section S12, Figure S1) explores this point further by showing how different hypothetical within-session decay rates would alter the predicted benefit of higher frequency at fixed weekly volume. Because these predictions are model-based rather than empirically estimated from the current data, they are provided as heuristic context rather than as additional evidence.

### 4.4. Pattern across contrasts and the ecological optimum

Reading the three contrasts together suggests a consistent pattern. For one versus two sessions per week, the five trials show similar hypertrophy at matched volume, with a pooled estimate slightly favoring the lower frequency and an interval too wide to support a clear advantage for either schedule; the trial most favoring the lower frequency (Gentil et al., 2018, in trained men) found significant thickness gains only in the once-weekly arm, and adding Tavares et al. (2017) in sensitivity analysis did not change the contrast. For one versus three sessions, the pooled estimate leans toward higher frequency (g = −0.23) but is non-significant; the lean comes chiefly from the within-subject MRI trial (Neves et al., 2022, −0.62, measured one week after training), with smaller contributions from one high-risk indirect-measure trial (McLester et al., 2000) and one direct-measure trial with imputed dispersion (Schoenfeld et al., 2015), and the four direct-measure trials pooled to −0.25 (95% CI −0.65 to 0.14). In Neves et al. (2022) the three-session leg, training at its own loads, accumulated 19.5% more volume load than the once-weekly leg, and the trial’s authors attribute the larger effect size in that condition to the extra volume load rather than to the schedule, since their volume-load-equated condition returned no difference (Section 3.3); the one-versus-three lean is therefore partly a volume-load effect that the sets-equated design does not remove. For two versus three sessions, the pooled effect is essentially zero, with one trial favoring the lower frequency (Lasevicius et al., 2019) and one the higher (Brigatto et al., 2022) by more than the trivial bound.

Within the studied range of one to three sessions and the volumes examined, the data do not show a material difference in hypertrophy from distributing an equated volume across one, two or three weekly sessions, and do not exclude a small one.

From a programming standpoint, the present data lack the precision to distinguish one, two or three weekly sessions at a fixed weekly volume. A default of two or three sessions per muscle per week is a programming convention supported by strength guidelines (Currier et al., 2026) and by practical considerations, such as session length and scheduling, that this review did not test. When weekly volume is high enough that a single weekly session would push per-session sets well above the range examined in Remmert et al. (2025), increasing frequency becomes a practical way to avoid very large sessions, but this scenario has not been directly tested in hypertrophy trials and should be treated as a hypothesis for future work rather than a confirmed effect.

Two further points from the trial set qualify this reading. First, the intervention durations of six to twelve weeks produced significant within-group hypertrophy in at least one arm in fourteen of the sixteen trials; the two indirect-measure one-versus-three trials detected no growth in either arm, so their between-group estimates near zero carry little information about a frequency effect, and the direct-measure one-versus-three estimate (Section 3.3) is the more informative figure for that contrast. A random-effects meta-regression treated duration as a modifier directly and found none that it could detect: the slope was small and non-significant (+0.02 g per week, 95% CI −0.08 to 0.12; p = 0.75), with the fitted estimate trivial at both ends of the range (approximately −0.10 at six weeks and −0.01 at twelve weeks) and a detectable slope of about 0.8 g across the range. The slope is too imprecise to say whether any lean changes with duration, and no inference about mechanism is drawn from its sign.

Second, the two-versus-three comparison was near zero in direct and indirect measures and in trained men, untrained mixed-sex and older or postmenopausal cohorts, although the older cohorts are two untrained trials and the inference across populations rests on them.

The relation to strength is less clear than is sometimes stated. All sixteen trials measured strength; fourteen reported no between-group difference, and two favored the higher frequency on one measure (McLester et al., 2000, leg press; Ochi et al., 2018, isometric strength in untrained men). Among the syntheses, Grgic et al. (2018) found no significant frequency effect on strength at equated volume (p = 0.42), Tao et al. (2026) reported an equal-volume strength estimate favoring higher frequency (g = −0.24), and Pelland et al. (2026) a volume-adjusted strength benefit of higher frequency with practice of the test among the candidate explanations. Evidence on strength at equated volume is therefore mixed, and the present review draws no inference about hypertrophy from it.

### 4.5. Scope and boundary conditions

#### High weekly volume

The high weekly-volume regime was not tested. Most included trials sat at moderate per-session volumes and so may have been unable to reveal a frequency benefit that exists at high weekly volumes, where a low-frequency schedule is forced past the productive per-session range. This defines the boundary of the claim, and it is identified as the priority for future work (Section 4.7).

Two observations bear on where that boundary lies. First, nine trials asked the lower-frequency arm to perform nine to eighteen sets for the measured muscle in one session (Gentil et al., 2015, 2018; Schoenfeld et al., 2015; Thomas & Burns, 2016; Neves et al., 2022; Calzavara & Barroso, 2026; Brigatto et al., 2019, 2022; Lasevicius et al., 2019), and none found a significant hypertrophy advantage for the higher frequency; Brigatto et al. (2019) reported effect sizes favoring the twice-weekly schedule, with load progression absent in the once-weekly arm, Brigatto et al. (2022), which equated eighteen weekly sets as nine versus six sets per session, reported small to moderate effect sizes favoring three sessions at three of four sites, and Neves et al. (2022) reported a moderate effect size favoring three sessions when both legs trained at their own loads (and none when volume load was held equal), so none of these trials shows the two frequencies as equivalent, but none found a significant difference.

Second, when the per-trial frequency effect was regressed on the lower-frequency arm’s per-session volume, and separately on weekly volume, neither moderated the outcome (per-session slope +0.024 g per set, p = 0.24; weekly slope +0.016, p = 0.15; Section 3.5 and Figure 4); both slopes are positive, the opposite of the direction a per-session ceiling requires, but the regressions could detect only a change of about 1.0 g across the observed range, so they do not exclude a smaller volume dependence. Within the tested range, therefore, the data do not show the within-session decay in per-set returns that a per-session ceiling would require, at the power available; little decay up to sixteen to eighteen sets is one of the explanations Pelland et al. (2026) offer for their flat frequency coefficient, and the present data neither confirm nor exclude it. They cannot speak to weekly volumes beyond those studied.

#### Measurement timing and swelling

A further boundary concerns measurement timing. Because the lower-frequency condition in a volume-equated design performs more sets in each session, any swelling that persists to the final measurement would inflate the lower-frequency arm and bias the standardized difference toward positive values, against the higher frequency. Residual swelling therefore cannot produce the absence of a frequency effect reported here; it could only conceal a small advantage of higher frequency.

Three observations limit that possibility. First, every direct-measure trial in the synthesis took its final measurement 48 hours or more after the last session (Section 3.3). Second, the number of sets in a session does not change the magnitude or time course of the swelling response in trained men (7, 14 or 21 sets, with thickness and echo intensity at baseline within 24 hours; Alvarez et al., 2026), although sessions of higher repetitions at moderate load can leave cross-sectional area modestly elevated at 48 hours (Bartolomei et al., 2017). Because the arms of each trial here differed in sets per session but used the same loads and repetition ranges (Section 2.2; in the condition of Neves et al., 2022, used here, both limbs trained at their own repetition-maximum loads), any residual offset at a 48 to 72 hour scan would be expected to be similar across arms and to cancel in the between-arm comparison; it would raise both arms’ pre-to-post changes rather than the difference between them. The converse direction also deserves stating: because the higher-frequency arm trains more often in the final week, swelling that persisted beyond 48 to 72 hours and accumulated across sessions could in principle inflate that arm instead; the return of thickness and echo intensity to baseline within 24 hours after 7, 14 or 21 sets in Alvarez et al. (2026) argues against accumulation in trained participants, but no included trial measured it.

Third, the setting in which swelling does distort size measures is the untrained state in the first weeks of training, where Damas et al. (2016) reported a 2.7% rise in vastus lateralis cross-sectional area accompanied by a 17.2% rise in echo intensity 72 hours after four sessions; the three untrained direct-measure trials here (Gentil et al., 2015; Ochi et al., 2018; Grzyb et al., 2022) measured at 48 hours to five days after eight to eleven weeks of training. In Damas et al. (2016), echo intensity was still elevated after 10 weeks, but the authors judged it comparatively minor against the 10.4% increase in cross-sectional area, and it was no longer elevated once normalized to it. Ochi et al. (2018), the trial in which a swelling bias would be largest (six sets against two per session, untrained men, 72 hours), returned a near-zero effect and found no between-group difference in muscle stiffness at the same time point.

The indirect-measure trials did not report the interval, and whole-body lean-mass estimates are in any case less sensitive to local fluid shifts; their pooled estimate (g = −0.04) did not differ from the direct-measure estimate (g = −0.07). The companion model-based analysis (Section 4.2) reached the same conclusion from the corpus side: its partitioning coefficient lay between −0.051 and −0.020 under five specifications that restrict or adjust for measurement timing.

#### Maintenance doses

Reduced-training studies suggest that previously accrued muscle can be preserved for extended periods on very low weekly doses, including a single weekly session of one set per exercise (about three sets per muscle in Bickel et al., 2011) in younger adults, provided relative load is maintained (Bickel et al., 2011; Spiering et al., 2021); older adults appear to need more (Spiering et al., 2021), and in Bickel et al. (2011) neither reduced dose maintained myofiber hypertrophy in the older group. Tavares et al. (2017), analyzed here in sensitivity analysis, is the one such study to compare frequencies at an equated reduced dose and found no difference between one and two sessions per week. Mpampoulis et al. (2024) varied the maintenance interval directly: after twelve weeks of concurrent training, young women who kept one session every seven days held quadriceps cross-sectional area over a further twelve weeks, whereas one session every fourteen days lost 5.9 ± 1.8% of cross-sectional area (53.8 to 50.7 cm²), about half the 5.8 cm² mean gain of the whole cohort during training (44.7 to 50.5 cm², n = 27; this subgroup’s own gain was not reported); the authors’ 94% is the share of post-training cross-sectional area kept, not of the gain. These findings bear on the minimum dose that preserves muscle rather than on how a growth-producing weekly volume is best distributed, and they do not alter the present conclusion. A more detailed discussion is provided in the Supplementary Material (Section S14).

#### Repetition range and load

Repetition range and relative load, examined as a further exploratory moderator, showed no relation to the frequency effect that the analysis could detect (Section 3.5, Figure 5). This coheres with the broader loading literature: across a wide span of the repetition continuum, load has little independent effect on hypertrophy once sets are taken near failure (Schoenfeld et al., 2021), so it would be surprising for load to govern how the resulting volume is best distributed.

The mechanistic rationale for expecting an interaction is nonetheless reasonable, since lower loads carried to failure produce greater acute neuromuscular and perceptual fatigue than heavier loads (Varela-Olalla et al., 2025) and greater discomfort (Fisher et al., 2017), and that fatigue changes motor unit recruitment across subsequent sets, with consequences for hypertrophy that remain unresolved (Flewwelling et al., 2025); a low-frequency schedule that compresses the weekly volume into one fatiguing session might therefore be expected to suffer disproportionately at higher repetitions. The present data neither confirm nor exclude that possibility, because the included trials were not designed to vary load: ten of sixteen sat at eight to twelve repetitions, so the contrast on which the hypothesis turns is barely represented. What can be said is that, within the moderate-load range that dominates the available frequency trials, no load dependence of the frequency effect large enough to be detected here was present.

### 4.6. Limitations

#### Size and precision of the evidence

The evidence base is small, comprising 16 pooled trials and 380 participants, with individual trials underpowered for between-group contrasts; the pooled analysis could itself detect a true effect of about 0.26 g with 80% power, and the optimal information size for the smallest effect of interest is about 785 participants. Two trials required imputation of change-score variance; the pooled estimate moved by 0.02 when they were removed. One trial (Gentil et al., 2018) was analyzed from its published participant-level data, and its effect, the largest in the pool, moved the pooled estimate by 0.03. The per-trial effect sizes for nine trials depend on an assumed pre-post correlation, and the close agreement among trials is partly a consequence of that assumption (Section 3.3).

#### Measurement

Most hypertrophy measures were site-specific and cannot speak to regional or whole-muscle growth; because sites were averaged within each trial, the review cannot examine the regional difference suggested by Tao et al. (2026) and Schoenfeld et al. (2019), and one trial contributed a single lower-body site because only that muscle was trained at an eligible contrast (Yue et al., 2018). Five trials used indirect lean-mass measures of limited sensitivity, two of them skinfold-derived, and none of the five reported the interval between the last training session and the post-intervention measurement. No included trial reported an index of muscle edema, so the absence of residual swelling at the final measurement is inferred from the measurement timing and from external data rather than measured directly. Several direct-measure trials measured at 48 to 72 hours, an interval at which a modest residual elevation of muscle size has been observed after high-repetition sessions in trained men (Bartolomei et al., 2017); within-trial comparisons are protected against a common offset, but absolute gains in either arm may be overstated.

Ten of the eleven pooled direct-measure trials assessed hypertrophy as ultrasound muscle thickness (A-mode in Brigatto et al., 2019, 2022; imaging ultrasound in the other eight); only Neves et al. (2022) used MRI. Thickness changes do not track MRI-measured changes reliably: in the largest comparison, vastus lateralis thickness captured under a third of the MRI-measured change and individual changes were unrelated (Balshaw et al., 2026), whereas in two smaller studies vastus lateralis thickness rose about as much as MRI-measured area (Franchi et al., 2018; Ruple et al., 2022; the differences were not tested). The pooled estimate therefore excludes large between-frequency differences in muscle thickness more firmly than it excludes differences in whole-muscle size.

#### Effort and achieved volume

Proximity to failure and exact per-session volumes were not standardized or fully reported across trials, so the stimulus was matched only at the level of prescribed sets. Equating sets does not equate achieved volume load: in the three trials that reported it, the higher-frequency arm lifted more volume load at the same relative load (16.3% more in Brigatto et al., 2019; 19.5% more in Neves et al., 2022; and more at each exercise in Calzavara & Barroso, 2026). Although eleven of the sixteen trials specified that all working sets reached muscular failure or volitional fatigue, only two reported effort on a repetitions-in-reserve scale, four prescribed load by repetition maximum or a percentage of one-repetition maximum without confirming failure, and one used a fixed submaximal load (Supplementary Table S1); in the volume-load-equated condition of Neves et al. (2022), used here in sensitivity analysis, the higher-frequency limb did not always reach failure, and that condition returned no difference between limbs (g = +0.04), whereas the sets-equated condition, in which both limbs trained to their own repetition maximum and the higher-frequency limb lifted more, returned g = −0.62 (Section 3.3). Because proximity to failure is likely to influence per-set hypertrophy and interacts with accumulated fatigue, in that a set carried to failure late in a large low-frequency session may fall short of one performed fresh at a higher frequency, this unmeasured variation in effort is a potential confounder of the frequency comparison, and of the volume-moderation analysis in particular.

#### Training history, duration and frequency range

In trained cohorts the frequency comparison is confounded with the novelty of the schedule: in Schoenfeld et al. (2015) and Brigatto et al. (2019) the higher-frequency arm was the less accustomed one, whereas in Gentil et al. (2018) the once-weekly arm was, and it alone gained thickness, so the confound does not run in one direction across the set. Interventions were short, from six to twelve weeks, so very long-term divergence cannot be excluded. The synthesis does not address frequencies above three sessions per muscle per week, nor the very high weekly-volume regime in which a frequency effect is most plausibly hypothesized; the one volume-equated trial reporting greater thickness gains for the higher frequency (Zaroni et al., 2019, one versus five sessions, with fifteen sets per muscle in one session in the once-weekly arm) lies outside the range by design.

#### Review process

Several procedural limitations also qualify the synthesis. Data extraction and risk-of-bias appraisal were carried out primarily by a single reviewer, title and abstract screening, the first assessment against the eligibility criteria and the extraction of the four added trials were performed with a generative AI tool and checked by the authors, and the review was not prospectively registered, its OSF project being created after the original searches and data extraction; these depart from the dual-independent, pre-registered process recommended for systematic reviews in the sport and exercise sciences (Ardern et al., 2022). To partially mitigate them, a second reviewer independently re-screened and re-extracted a subsample of trials (Section 3.1), all sixteen pooled effect sizes were recomputed from the trial reports in code separate from the authors’ (a check of computation rather than of extraction for the four trials the AI tool extracted), and the screening log, data and code are openly deposited, with the analyses not named in the OSF project description identified in Section 2.1.

#### Search

The searches were re-run in full in October 2026 because complete screening records of the April 2026 searches had not been retained. Five of the sixteen trials were not retrieved by the database strings and were found in reference lists or the earlier search, which suggests that the strings miss trials of this design described in other terms (for example “workout frequency” or “times a week”) and that further trials may have been missed. The reference list of one review could be read but not its table of included studies (Kneffel et al., 2021). Finally, the search was restricted to English-language reports, and no trial registry or gray-literature source other than CENTRAL was searched, so language and publication-selection biases cannot be excluded, although the funnel and contour-enhanced plots showed no evidence of small-study effects.

### 4.7. Directions for future research

The priority is a trial conducted at high weekly volume, where a low-frequency schedule would be forced past the productive per-session ceiling, since this is the single regime in which a hypertrophic frequency effect is most plausible and the present trials are silent. Such trials should sample hypertrophy regionally, using multiple sites or full muscle-length magnetic-resonance or computed-tomography panels rather than a single slice, so that regional growth cannot be averaged away. They should measure and report proximity to failure, and equate arms on measured proximity to failure rather than on prescribed sets alone, since ‘effective’ or ‘stimulating’ repetitions have no validated measure. Trials in trained participants should counterbalance which schedule is the unaccustomed one.

Longer interventions and frequencies above three sessions per muscle per week would extend the dose-response beyond the range examined here, and trials with a sample size approaching the optimal information size for a small effect (about 785 participants in total, or fewer with a within-subject design and a direct measure) would resolve the small effects that the present pool cannot. Until such evidence exists, the present conclusion should be read as bounded to the volume range that the available trials occupy.

### 4.8. Practical applications

Within the range of one to three sessions per muscle per week and the weekly volumes studied, the present findings support placing weekly volume ahead of frequency in hypertrophy-oriented programming decisions. In practice, coaches and practitioners can first establish a target number of hard sets per muscle per week, then distribute that volume across one, two or three weekly sessions in whichever pattern best fits the athlete’s schedule, recovery capacity and tolerance for per-session volume, with the reservation that a small advantage of the higher frequency, of the order the confidence interval admits, cannot be excluded. The present data lack the precision to distinguish one, two or three sessions at a fixed weekly volume; a default of two or three sessions per muscle per week is a programming convention supported by strength guidelines (Currier et al., 2026) and by practical considerations that this review did not test.

Higher frequencies are most likely to be useful where weekly volume is sufficiently high that fewer sessions would necessitate very large, fatiguing workouts, or where strength and skill are co-primary goals; the strength advantage of higher frequency is smaller when volume is equated (Currier et al., 2026) and the evidence for it at equated volume is mixed (Section 4.4).

## 5. Conclusions

This systematic review and meta-analysis show that, within the ecologically common range of one to three sessions per muscle per week and the weekly volumes studied, resistance training frequency has no independent influence on hypertrophy that the available trials could detect when weekly volume is equated; a small effect favoring the higher frequency, and for one versus three sessions a moderate one, is not excluded. The pooled standardized mean difference across sixteen trials was trivial and non-significant (g = −0.06, 95% CI −0.24 to 0.13); the contrast-specific estimates for 1 vs 2 and 2 vs 3 sessions per week were trivial and the 1 vs 3 estimate small and non-significant (g = −0.23, 95% CI −0.57 to 0.10), with no moderation by intervention duration, weekly or per-session set volume, repetition range or relative load that the analyses could detect. Although the certainty of evidence was rated low to very low, reflecting small samples and short interventions, the convergence of point estimates across direct and indirect hypertrophy measures and across trained and untrained cohorts indicates that large, practically important hypertrophic effects of altering frequency at equated volume are unlikely in typical programs, with the qualification that most trials measured muscle thickness, which does not track whole-muscle growth reliably (Section 4.6); because ten of the eleven direct-measure trials used ultrasound thickness, the pool excludes large between-frequency differences in thickness more firmly than it excludes differences in whole-muscle size.

For practitioners, these findings support treating weekly volume per muscle as the primary focus when designing hypertrophy-oriented resistance training, with frequency used mainly as a lever to distribute that volume, manage per-session load and support adherence and recovery. The data lack the precision to distinguish one, two or three sessions at a fixed weekly volume; a default of two or three sessions per muscle per week is a convention supported by strength guidelines and practical considerations rather than a finding of this review. Future work extending interventions to longer durations, testing higher weekly volumes and frequencies beyond three sessions per week, measuring proximity to failure, and assessing hypertrophy regionally with whole-muscle imaging will determine whether the small effects that the present evidence cannot exclude are real.

## Supplemental material

Tables S1 to S7 (effort and proximity-to-failure prescription; exclusions at eligibility assessment, with reasons; records identified by database; moderator values and counting rule; the Brigatto et al. (2022) composite under alternative dispersion sources; contrast-level intervals under alternative conventions; the cross-check against Tao et al., 2026), Supplementary Methods S1 to S11 and Notes S12 to S14, and Figures S1 and S2 are available online.

## Ethics statement

This study is a systematic review and meta-analysis of previously published data and did not involve new data collection from human participants; institutional review board approval and informed consent were therefore not applicable.

## Declarations

### Author contributions

A.G.B. conceived and designed the review, performed the analysis, curated the data, and wrote the original draft. M.M. independently re-screened and re-extracted a subsample of trials and re-extracted the four trials added in the October 2026 update (validation). Both authors reviewed and edited the manuscript and approved the submitted version.

### Declaration of generative AI use

The authors used the generative AI tool Anthropic Claude during the October 2026 update of this review. Under the authors’ direction, it ran the database searches, performed title and abstract screening, assessed records against the eligibility criteria, extracted data from the newly included trials, and recomputed the effect sizes and meta-analyses in code separate from the authors’ own. Every decision at eligibility assessment and every extracted value was checked by the authors. M.M. independently re-extracted the four added trials. It was also used to produce figures and tables. Both authors confirm that they understand and have verified the analyses conducted, including the effect-size calculations, the meta-analyses and the code that produced them, and both take full responsibility for the content of the manuscript and of the materials, including the Supplementary Material and the data, code and screening records deposited on the Open Science Framework.

## Data availability statement

The screening log, extracted data, effect-size calculations, and analysis code supporting the findings of this study are openly available on the Open Science Framework (OSF) at https://osf.io/ebrcx.

