## Supplementary Material for "Resistance Training Frequency and Muscle Hypertrophy: A Systematic Review and Meta-Analysis of Volume-Equated Trials"

**Contents**

Supplementary Methods S1 to S11; Supplementary Notes S12 to S14; Supplementary Tables S1 to S7 (Table S7 appears within Note S13); Supplementary Figures S1 and S2.

**Supplementary Methods**

**S1. Database search strategy**

Five electronic databases were searched on 6 October 2026, through institutional library access: MEDLINE through PubMed, Scopus, Web of Science Core Collection, SPORTDiscus with Full Text through EBSCO, and the Cochrane Central Register of Controlled Trials (CENTRAL) through Ovid EBM Reviews (August 2026 issue). These searches replace searches of the same five databases run in April 2026, for which complete screening records were not retained. The strategy was structured around three conceptual domains combined with AND, Domain A: resistance or strength training (intervention); Domain B: training frequency (variable of interest); Domain C: muscle hypertrophy or size (outcome), with synonymous terms within each domain combined with OR, and phrase searching with quotation marks. Records were limited to English. There was no date limit. The number of records from each database is given in Table S3, and each export is deposited with the screening log.

**S1.1 MEDLINE/PubMed, searched 6 October 2026**

((“Resistance Training”[MeSH Terms] OR “resistance training”[tiab] OR “strength training”[tiab] OR “weight training”[tiab] OR “weightlifting”[tiab] OR “weight lifting”[tiab] OR “progressive resistance training”[tiab] OR “progressive overload”[tiab] OR “RT”[tiab] OR “exercise training”[tiab]) AND (“training frequency”[tiab] OR “exercise frequency”[tiab] OR “session frequency”[tiab] OR “sessions per week”[tiab] OR “times per week”[tiab] OR “weekly frequency”[tiab] OR “frequency of training”[tiab] OR “training sessions”[tiab] OR “once per week”[tiab] OR “twice per week”[tiab] OR “three times per week”[tiab] OR “1x per week”[tiab] OR “2x per week”[tiab] OR “3x per week”[tiab] OR “daily training”[tiab] OR “split routine”[tiab] OR “full body”[tiab] OR “body part split”[tiab]) AND (“Muscle, Skeletal”[MeSH Terms] OR “Hypertrophy”[MeSH Terms] OR “muscle hypertrophy”[tiab] OR “muscle size”[tiab] OR “muscle mass”[tiab] OR “muscle thickness”[tiab] OR “cross-sectional area”[tiab] OR “CSA”[tiab] OR “lean mass”[tiab] OR “lean body mass”[tiab] OR “fat-free mass”[tiab] OR “muscle growth”[tiab] OR “hypertrophic”[tiab] OR “skeletal muscle”[tiab] OR “ultrasonography”[tiab] OR “ultrasound”[tiab] OR “MRI”[tiab] OR “DXA”[tiab] OR “DEXA”[tiab]))

*Filters: Humans; English; Randomized Controlled Trial, Controlled Clinical Trial, Clinical Trial. No date limit. 709 records.*

**S1.2 Scopus (Elsevier), searched 6 October 2026**

TITLE-ABS-KEY((“resistance training” OR “strength training” OR “weight training” OR “weightlifting” OR “progressive resistance” OR “exercise training”) AND (“training frequency” OR “exercise frequency” OR “sessions per week” OR “times per week” OR “weekly frequency” OR “session frequency” OR “once per week” OR “twice per week” OR “three times per week” OR “split routine” OR “full body training” OR “body part split”) AND (“muscle hypertrophy” OR “muscle size” OR “muscle mass” OR “muscle thickness” OR “cross-sectional area” OR “lean mass” OR “lean body mass” OR “fat-free mass” OR “muscle growth” OR “hypertrophic adaptation” OR “skeletal muscle” OR “ultrasound” OR “ultrasonography” OR “MRI” OR “DXA” OR “DEXA”)) AND LANGUAGE(english) AND DOCTYPE(ar OR re)

*Document types: Article (ar), Review (re). Language: English. No date limit. 1,178 records.*

**S1.3 Web of Science Core Collection (Clarivate), searched 6 October 2026**

TS=(“resistance training” OR “strength training” OR “weight training” OR “weightlifting” OR “progressive resistance training”) AND TS=(“training frequency” OR “exercise frequency” OR “sessions per week” OR “times per week” OR “weekly frequency” OR “once per week” OR “twice per week” OR “three times per week” OR “split routine” OR “full body training”) AND TS=(“muscle hypertrophy” OR “muscle size” OR “muscle mass” OR “muscle thickness” OR “cross-sectional area” OR “lean mass” OR “fat-free mass” OR “muscle growth” OR “hypertrophic” OR “ultrasound” OR “ultrasonography” OR “MRI” OR “DXA”)

*Refined by: Language = English; Document Types = Article OR Review. No date limit. 567 records (582 before the limits).*

**S1.4 SPORTDiscus with Full Text (EBSCO), searched 6 October 2026**

( DE “RESISTANCE training” OR DE “WEIGHT training” OR DE “STRENGTH training” OR TI (“resistance training” OR “strength training” OR “weight training” OR “progressive resistance” OR “exercise training”) OR AB (“resistance training” OR “strength training” OR “weight training” OR “progressive resistance” OR “exercise training”) ) AND ( TI (“training frequency” OR “exercise frequency” OR “sessions per week” OR “times per week” OR “weekly frequency” OR “once weekly” OR “twice weekly” OR “three times weekly” OR “split routine” OR “full body” OR “frequency of resistance training”) OR AB (“training frequency” OR “exercise frequency” OR “sessions per week” OR “times per week” OR “weekly frequency” OR “once weekly” OR “twice weekly” OR “three times weekly” OR “split routine” OR “full body” OR “frequency of resistance training”) ) AND ( DE “MUSCLE hypertrophy” OR DE “MUSCLES – Hypertrophy” OR TI (“muscle hypertrophy” OR “muscle size” OR “muscle mass” OR “muscle thickness” OR “cross-sectional area” OR “lean mass” OR “fat-free mass” OR “muscle growth” OR “hypertrophic” OR “ultrasonography” OR “MRI” OR “DXA” OR “DEXA”) OR AB (“muscle hypertrophy” OR “muscle size” OR “muscle mass” OR “muscle thickness” OR “cross-sectional area” OR “lean mass” OR “fat-free mass” OR “muscle growth” OR “hypertrophic” OR “ultrasonography” OR “MRI” OR “DXA” OR “DEXA”) )

*Boolean search mode. Limiters: Peer Reviewed; Language = English. No date limit. 378 records (403 before the limiters). Each domain is enclosed in parentheses so that the three domains combine with AND as intended.*

**S1.5 Cochrane CENTRAL (Ovid EBM Reviews, August 2026 issue), searched 6 October 2026**

1 exp Resistance Training/ 2 (“resistance training” or “strength training” or “weight training” or “progressive resistance”).ti,ab,kw. 3 (“training frequency” or “sessions per week” or “times per week” or “weekly frequency” or “once per week” or “twice per week” or “three times per week” or “split routine” or “full body”).ti,ab,kw. 4 exp Hypertrophy/ 5 exp Muscle, Skeletal/ 6 (“muscle hypertrophy” or “muscle size” or “muscle mass” or “muscle thickness” or “cross-sectional area” or “lean mass” or “fat-free mass”).ti,ab,kw. 7 1 or 2 8 4 or 5 or 6 9 3 and 7 and 8 10 limit 9 to english language

*779 records at line 9; 773 after the English limit. The Cochrane Library interface was not used.*

**S1.6 Supplementary searches**

The reference lists of seven frequency reviews were searched by hand: Cuthbert et al. (2021), Grgic et al. (2018), Kneffel et al. (2021), Ramos-Campo et al. (2024), Schoenfeld et al. (2016), Schoenfeld et al. (2019) and Tao et al. (2026). For Kneffel et al. (2021), only the reference list could be read, not the table of included studies. These lists yielded nine reports for assessment, of which four were included (Gentil et al., 2015; Grzyb et al., 2022; Prieto González et al., 2020; Yue et al., 2018). One further trial (Brigatto et al., 2022), identified in the April 2026 search and not retrieved by the October 2026 strings, was assessed and included. Five of the sixteen pooled trials were therefore found outside the database searches. No trial registry beyond CENTRAL and no gray-literature source was searched, and no forward citation search was run for the October 2026 update.

**S2. Inclusion and exclusion criteria**

Trials were eligible if they (i) used a longitudinal resistance-training intervention of at least six weeks; (ii) compared, for the measured muscle, training frequencies of one, two or three sessions per week, so that the eligible contrasts were one versus two, one versus three and two versus three, and a comparison in which the measured muscle was trained more than three times per week in any arm was ineligible; (iii) assigned the frequencies by the investigators rather than by self-selection or attendance; (iv) equated total weekly volume between conditions, with the per-session set count adjusted to hold weekly sets, repetitions, and load constant; and (v) reported a direct or indirect measure of muscle size (ultrasound or MRI muscle thickness or cross-sectional area, or an estimate of lean mass by DXA or anthropometry, including skinfold-derived lean mass).

Bioelectrical impedance was not accepted; one trial was excluded on that ground (Cardoso et al., 2024), and its split arm trained each muscle 1.5 times per week on average, outside the eligible contrasts. Where a trial compared eligible frequencies for some measured muscles and not others, only those muscles were used: in Yue et al. (2018), only the vastus medialis, trained once versus twice weekly. Designs that fixed per-session sets and therefore allowed weekly volume to differ between frequencies were excluded, because they confound frequency with volume.

One trial (Tavares et al., 2017) compared one with two sessions per week with sets equated (four sets of each exercise once weekly against two sets twice weekly) and volume-load described by its authors as equalized, during an eight-week reduced-training phase at about half the preceding volume-load that followed an eight-week standardized program; because neither arm gained muscle size and the quantity measured is retention under a reduced dose, it was analyzed in sensitivity analysis only (main text Section 3.3).

Throughout, hypertrophy is the umbrella construct (an increase in muscle size). Its operationalizations are distinguished: muscle size denotes direct, site-specific measures (ultrasound or MRI thickness or CSA), whereas lean or muscle mass denotes indirect whole-body or regional estimates (DXA or skinfold-derived). Because the indirect measures are less sensitive to localized growth, this distinction is carried through the analysis and its limitations.

**S3. Study selection and screening**

Records from the five databases were merged and deduplicated in three passes: by DOI, by exact match of the normalized title, and by near-identical title (similarity of 96% or more) with the same publication year. Titles and abstracts were then screened against the eligibility criteria. Every record not excluded at that stage was assessed against the full criteria, on the full text where the abstract left any criterion open and on the abstract alone where it stated the failing design feature explicitly (for example, an arm above three sessions per muscle per week, or the same per-session prescription in every arm). Each decision, its reason and the basis on which it was made (title and abstract, abstract, or full text) are recorded in the screening log deposited on OSF; Table S2 lists the exclusions at eligibility assessment.

Title and abstract screening and the first assessment against the eligibility criteria were performed with a generative AI tool under the authors' direction, and the authors checked every decision at eligibility assessment and a random sample of 100 of the 1,823 title and abstract exclusions other than reviews (seed 20261007); all 79 eligibility decisions and all 100 sampled exclusions were confirmed.

For the original twelve trials, data extraction was performed primarily by a single reviewer; for the four trials added to the pool in October 2026, data were extracted with the generative AI tool and checked against the trial reports by the authors. To assess accuracy, a second reviewer independently re-screened and re-extracted seven trials of the original set: five randomly selected trials and the two trials flagged at screening in the original set for absent dispersion data (Gentil et al., 2018, before its participant-level data were retrieved; Schoenfeld et al., 2015), with random seeds documented for reproducibility; Candow and Burke (2007), whose dispersion was also imputed, fell outside the random draw. The second reviewer also re-extracted the four trials added in the October 2026 update and agreed with every coded design item and extracted value checked; the participant-level changes for Gentil et al. (2018) were computed from the trial's published data file and were not recomputed by him. Percentage agreement was calculated for eligibility decisions and categorical extraction fields; Cohen's kappa is undefined here because every re-screened trial was an include, so expected agreement equals observed agreement. Agreement is reported in Results Section 3.1.

All sixteen pooled effect sizes were also recomputed from the trial reports with the same tool, under the dispersion rule in S4, in code separate from the authors' own; for the four added trials this checks the computation but not the extraction.

**S4. Data extraction and coding**

For each trial we extracted, per group and per measured site, the pre- and post-intervention means and standard deviations, the sample size, the per-muscle frequency contrast, the intervention duration, the training status of the sample, the hypertrophy measurement method, the interval between the last training session and the post-intervention measurement, and whether any index of muscle edema was reported.

One rule was applied to dispersion. Where a trial reported change-score dispersion directly, those values were used: change-score standard deviations for Neves et al. (2022), Thomas and Burns (2016) and Yue et al. (2018); standard deviations recovered from the 95% confidence intervals of the mean change for Grzyb et al. (2022), where the interval printed for the knee-extensor change in the three-session group (0.31, 0.09 to 0.12 cm) does not contain its point estimate and was reconstructed symmetrically from its lower bound (0.09 to 0.53 cm; the composite is -0.21 under that reconstruction and -0.24 if the two-session group's standard deviation is used for that site instead); and the trial-reported between-group standardized differences of the changes for Brigatto et al. (2022) (Table S5). Pina et al. (2020) reported covariate-adjusted mean changes with confidence intervals; because the two groups' changes were equal, its effect size is 0.00 under either source.

Where only pre and post standard deviations were reported, the change-score standard deviation was derived with an assumed pre-post correlation (S5). For Schoenfeld et al. (2015), which presents means with standard deviations in figures, and Candow and Burke (2007), which reports baseline standard errors in a table and post-intervention values in a figure, the standard deviations were read from the figures and the derived change-score standard deviations are treated as imputed, flagged and tested in sensitivity analysis.

Gentil et al. (2018) presents muscle thickness only as an individual-values figure; its effect size was computed from the participant-level data published with the trial (once weekly, change 1.14 ± 0.87 mm; twice weekly, 0.41 ± 0.93 mm; n = 8 per group), which give each participant's change directly, so no correlation was assumed. For Yue et al. (2018), the change in the twice-weekly arm is taken from its Table 3 (1.8 ± 2.43 mm); the Abstract prints 1.82, which gives g = +0.16 rather than +0.17. In the submitted version of this review the supplementary file had not been retrieved and the trial was synthesized narratively.

**S5. Effect-size calculation**

The effect measure was the change-score standardized mean difference (Hedges g), computed as the between-group difference in mean change divided by the pooled change-score standard deviation, with the small-sample correction applied (Hedges, 1981). The sign convention was set so that a negative g favors the higher-frequency condition. A smallest effect size of interest was set at $|g| = 0.2$, the conventional boundary between a trivial and a small standardized effect; pooled point estimates below this bound are interpreted as trivial, and only intervals contained within it as excluding even a small effect. Where only pre and post standard deviations were reported, change-score standard deviations were derived from them using a pre-post correlation of $r = 0.7$, with sensitivity analyses at $r = 0.5$ and $r = 0.9$. Each such effect size scales by the square root of 0.6 divided by 2(1 - r), by 0.77 at $r = 0.5$ and 1.73 at $r = 0.9$.

**S6. Multi-site aggregation**

Trials reporting multiple sites were aggregated to a single composite effect using the dependent-effects formula of Borenstein et al. (2009), assuming a within-study correlation among sites of 0.6. The correlation enters the composite through its sampling variance and therefore through the trial's weight in the pool: a four-site composite carries about 43% more weight at a correlation of 0.6 than at 1.0. The pooled estimate was recomputed at correlations of 0.3, 0.8 and 1.0 (main text Section 3.3).

**S7. Meta-analytic model**

Effects were pooled with a random-effects model with the between-trial variance estimated by restricted maximum likelihood; where that estimate is zero, as it was in every model, the estimate coincides with the DerSimonian and Laird (1986) estimate. Confidence intervals are reported with one convention throughout: the Wald interval from the inverse-variance weights. The Hartung-Knapp-Sidik-Jonkman (HKSJ) adjustment scales the standard error by the observed dispersion of the trials around the pooled estimate; when the trials agree more closely than chance would produce, the unmodified adjustment narrows the interval below the Wald interval (here to -0.21 to 0.09 for the overall estimate), and the standard remedy is to truncate the scaling factor at one, which gives -0.26 to 0.14. Table S6 gives the Wald, truncated HKSJ and unmodified HKSJ intervals for the overall estimate and each contrast.

**S8. Heterogeneity and prediction interval**

Heterogeneity was summarized with $I^2$ (Higgins et al., 2003), $\tau^2$, and a prediction interval. The overall pool returned $Q = 8.45$ on 15 degrees of freedom ($p = 0.90$), $I^2 = 0\%$ and $\tau^2 = 0$, and a prediction interval of -0.26 to 0.14; with $\tau^2$ at its boundary the prediction interval is close to the confidence interval, and the boundary estimate is interpreted cautiously given the small number of trials.

**S9. Sensitivity and subgroup analyses**

The stability of the pooled estimate was examined by leave-one-out analysis, by restricting to direct measures of muscle size, by removing trials with imputed variance, by removing the trials at high risk of bias, by restricting to trials whose final measurement was taken 48 hours or more after the last training session, by training status, by removing the four trials added in the October 2026 update, and by adding Tavares et al. (2017).

Intervention length was tested as a moderator by regressing the change-score effects on trial duration (weeks) in a random-effects (method-of-moments) meta-regression, reporting the slope, its 95% confidence interval, and residual heterogeneity. Further moderator analyses (per-session and weekly set volume, mean repetitions, and relative load) were exploratory and interpretively secondary; with at most 16 trials per model and no correction for multiple testing, they are reported as hypothesis-generating, each with the slope detectable at 80% power (2.8 standard errors). The moderator values and the counting rule are given in Table S4.

The within-subject trial (Neves et al., 2022) ran two conditions in separate groups of twelve men: in one both legs trained at their own repetition-maximum loads for the same nine weekly sets (sets equated, with the three-session leg accumulating 19.5% more volume load, 164,894 against 137,986 kg), and in the other the three-session leg was held to the once-weekly leg's mean load per repetition (volume load also equated, with sets that the authors note were not always maximal). The sets-equated condition is used (g = -0.62, 95% CI -1.44 to 0.20, from the reported change scores of 1.4 ± 3.0 and 3.9 ± 4.6 $cm^2$), because it matches the prescription rule of every other trial in the pool and the coding of the trial in the corpus analyzed by the companion paper; the first version of this review used the volume-load-equated condition (g = +0.04, 95% CI -0.76 to 0.84), which is retained as a sensitivity analysis in Section 3.3 of the main text. Because the trial reported contralateral limbs without a within-limb correlation, its variance was computed with the independent-groups standardized-mean-difference formula, a conservative choice that does not exploit the smaller standard error a paired analysis would yield.

**S10. Small-study effects and certainty of evidence**

Small-study effects were assessed with Egger's regression using both a standard-error moderator (intercept +0.28, SE 1.12, $t(14) = 0.25$, $p = 0.81$) and a sample-size-based ($1/\sqrt{n}$) precision term (intercept +0.44, SE 2.35, $t(14) = 0.18$, $p = 0.86$); the two agree. The second is reported because the standardized mean difference is algebraically correlated with its own standard error, so an asymmetry on the SE moderator alone can be an artifact of the effect-size metric rather than evidence of small-study bias. Risk of bias was appraised with the Cochrane RoB 2 tool (Sterne et al., 2019) across its five domains, supported where relevant by PEDro-scale criteria (de Morton, 2009); appraisal was single-reviewer. The certainty of the pooled estimate was rated with GRADE (Guyatt et al., 2008). The principal sources of bias in this literature are small samples, short interventions, unblinded outcome assessment, per-protocol analysis after exclusions for non-adherence and, in two trials, incompletely reported dispersion.

**S11. Software and reproducibility**

Analyses were performed in Python 3.11 (NumPy 1.26 and SciPy 1.11). The pooled estimate, its Wald and unmodified HKSJ intervals, $\tau^2$, $I^2$, Q, the three contrast estimates, the duration meta-regression and Egger's test were reproduced in the R package metafor 4.4 (Viechtbauer, 2010; *Journal of Statistical Software*, 36(3), 1–48) as an independent cross-check, to the fourth decimal place. The prediction interval reported here uses a t distribution on k - 2 degrees of freedom (Higgins, Thompson & Spiegelhalter, 2009; *Journal of the Royal Statistical Society: Series A*, 172, 137–159), so it is slightly wider than metafor's normal-based default, which coincides with the confidence interval when $\tau^2 = 0$. The code, the extracted data, the effect-size calculations, the database exports and the screening log are deposited on the Open Science Framework https://osf.io/ebrcx.

### Supplementary Notes

#### S12. Geometric-decay illustration

At a fixed weekly volume, a benefit of higher frequency requires that stimulus per set decline within a session, so that spreading the same weekly sets across more sessions recovers stimulus that would otherwise be lost to within-session decay. Figure S1 illustrates this dependence: it plots the hypertrophy advantage that a simple geometric within-session decay model would predict for a higher-frequency schedule (three versus one session per week) at fixed weekly volumes, as a function of the assumed per-set decay rate. The predictions are model-based rather than estimated from the present data and are provided as heuristic context only; at zero within-session decay the model predicts no frequency benefit, consistent with the trivial pooled effect observed here.

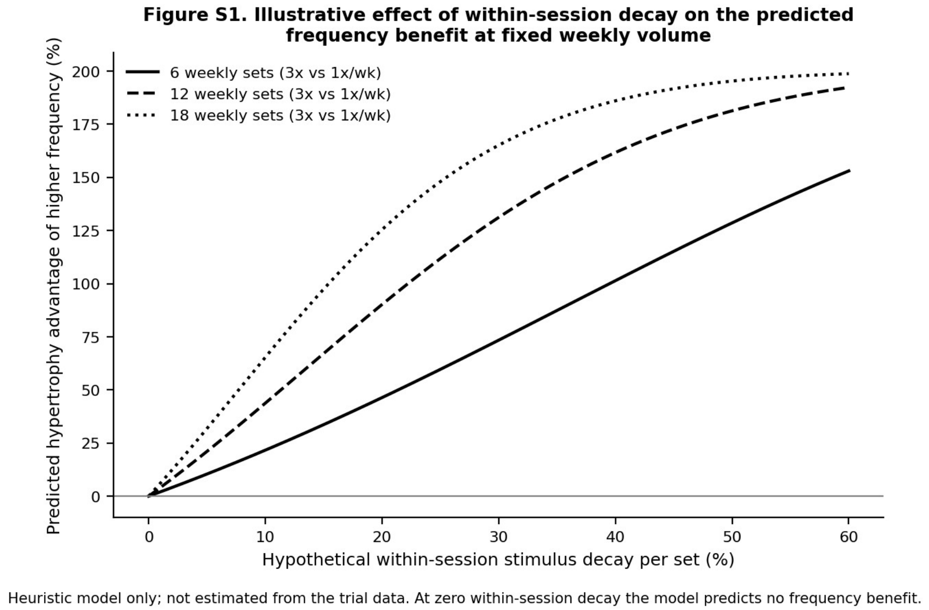


***Figure S1.*** *Illustrative effect of hypothetical within-session stimulus decay on the predicted hypertrophy advantage of higher training frequency at fixed weekly volume (heuristic model; not estimated from the trial data).*

#### S13. Cross-check against Tao et al. (2026)

The most directly comparable recent synthesis is Tao et al. (2026), which reached a similar conclusion for muscle mass in volume-equated comparisons (equal-volume subgroup g = 0.01) despite different analytic choices. Their approach to deriving change-score dispersion and to coding frequency contrasts yielded substantially larger per-trial standardized mean differences and higher apparent heterogeneity than the present extraction. The two syntheses therefore agree on the substantive conclusion, no meaningful independent effect of frequency on muscle size at equated volume, while differing in the magnitude and dispersion of the per-trial estimates, a difference attributable to methodological choices rather than to conflicting underlying data.

Table S7 sets the present review's per-trial composites beside the per-outcome effects Tao et al. extracted from the same trials, for the six trials of the original set common to both syntheses.

**Table S7. Per-trial composites of the present review beside the per-outcome effects extracted by Tao et al. (2026)**

| Trial | Contrast (here) | Our composite g [95% CI] | Tao extracted effects (n; g range; mean) | Note |
|---|---|---|---|---|
| Gentil (2015) | 1 vs 2 | -0.20 [-0.92, 0.52] | 2 effects; -0.95 to -0.78 (mean -0.86) | separate outcomes not aggregated |
| Brigatto (2019) | 1 vs 2 | -0.06 [-0.79, 0.68] | 4 effects; -0.86 to +0.09 (mean -0.25) | four measurement sites kept separate |
| Ochi (2018) | 1 vs 3 | -0.03 [-0.77, 0.70] | 7 effects; -0.74 to +1.52 (mean +0.02) | wide spread across measures/timepoints |
| Neves (2022) | 1 vs 3 | -0.62 [-1.44, 0.20] | 2 effects; -0.96 to +0.06 (mean -0.45) | Tao enters both conditions as separate effects; the composite here is the sets-equated condition |
| Lasevicius (2019) | 2 vs 3 | +0.42 [-0.20, 1.05] | 4 effects; -2.05 to -0.99 (mean -1.62) | Tao codes the contrast as 2v4 (weekly sessions), not 2v3 per muscle |
| Pina (2020) | 2 vs 3 | +0.00 [-0.57, 0.57] | 3 effects; -1.58 to 0.00 (mean -0.94) | separate sites/outcomes kept separate |

*Our composite g is from Table 2 of the main text (negative favors higher frequency). Tao values are the individual muscle-mass effect sizes in the openly deposited Tao et al. extraction; n is the number of separate effects they retained for that trial.*

Across these six shared trials, the present review's conservative per-trial composites cluster near zero (|g| ≤ 0.62), whereas Tao et al.'s per-outcome extractions span |g| up to roughly two and diverge in sign within a single trial (for example, Ochi 2018 ranges from -0.74 to +1.52), which mechanically inflates apparent between-trial heterogeneity. Two extraction choices account for the difference: Tao retains multiple site- and timepoint-specific effects per trial rather than aggregating them into a single composite with a conservative dependent-effects variance; and Tao codes frequency by total weekly sessions (coding Lasevicius 2019 as 2v4) rather than by per-muscle frequency (2v3). Both choices enlarge the per-trial standardized mean differences and their scatter. Despite this, the two syntheses converge on the same substantive result for volume-equated muscle mass: Tao's equal-volume subgroup pools to g = 0.01 and the present review to g = -0.06. The divergence is therefore one of extraction and contrast-coding, not of the underlying trial data. Tao et al. also reported a secondary signal favoring higher frequency for upper-body muscle; because the present composites average sites within each trial, region cannot be examined here.

#### S14. Maintenance-dose data and the current findings

Reduced-training studies indicate that previously accrued muscle can be preserved for extended periods on very low weekly doses. In Bickel et al. (2011), after 16 weeks of training three times per week, younger adults maintained myofiber hypertrophy for 32 weeks on one weekly session of either three sets per exercise (about nine weekly sets to the quadriceps) or one set per exercise (about three), with relative load maintained; neither reduced dose maintained myofiber hypertrophy in the older adults, and thigh lean mass by dual-energy X-ray absorptiometry fell on the lower dose. Spiering et al. (2021) summarize the minimal dose for younger populations as one session per week of one set per exercise and note that older populations may need up to two sessions per week and two to three sets per exercise.

Tavares et al. (2017), analyzed in the main text in sensitivity analysis, compared one and two weekly sessions with sets equated at a reduced volume-load after an eight-week standardized program and found no difference in quadriceps cross-sectional area retention. Mpampoulis et al. (2024) varied the interval itself: after twelve weeks of concurrent resistance and aerobic training, young women assigned to one session every seven days held leg-press strength and quadriceps cross-sectional area over a further twelve weeks, whereas those assigned to one session every fourteen days lost 5.9 ± 1.8% of cross-sectional area (53.8 to 50.7 cm²), about half the 5.8 cm² mean gain of the whole cohort during training (44.7 to 50.5 cm², n = 27; this subgroup's own gain was not reported; the authors' 94% is the share of post-training cross-sectional area kept, not of the gain); a detrained group returned to pre-training values. These findings show that preserving existing tissue requires far less weekly stimulus than adding to it. They bear on the minimum effective dose rather than on how a growth-producing weekly volume is best distributed across sessions, which is the question addressed here; two of the sixteen pooled trials are in older women, so the age dependence of the maintenance dose is a reason not to extend the maintenance findings to them.

### Supplementary Tables

#### Table S1. Effort and proximity-to-failure prescription in the included trials

| Trial | Contrast | Effort / proximity-to-failure prescription | Source |
|---|---|---|---|
| Gentil et al. (2015) | 1 vs 2 | 3 sets of 8–12 maximum repetitions; all sets performed to concentric failure (verbally encouraged). | Methods, RT intervention |
| Brigatto et al. (2019) | 1 vs 2 | 8–12 RM; all sets to momentary muscular failure (RPE/RIR of 9.5–10 reported for all sets). 17 of 20 participants had trained the lower body once weekly before the trial. | Methods, training protocol |
| Gentil et al. (2018) | 1 vs 2 | 3 sets of 8–12 repetitions to momentary concentric failure; loads adjusted to maintain the rep range. | Methods, RT intervention |
| Yue et al. (2018) | 1 vs 2 | 8 to 12 self-determined maximum repetitions at about 75% 1RM; up to 30 s rest within a set allowed to reach 8 repetitions; load raised when more than 12 were possible. Failure not stated. | Methods, Training intervention |
| Prieto González et al. (2020) | 1 vs 2 | 3 sets of 6 to 12 repetitions per exercise, intensity rising from 62% to 78% of estimated maximum; failure not stated. | Methods |
| McLester | 1 vs | Periodized repetition range 3–10 (≈80% 1RM start); every set to momentary muscular | Methods, |

| Trial | Contrast | Effort / proximity-to-failure prescription | Source |
|---|---|---|---|
| et al. (2000) | 3 | failure. | Experimental Design |
| Schoenfeld et al. (2015) | 1 vs 3 | 2 to 3 sets per exercise of 8–12 RM (18 sets per session); each set to momentary concentric muscular failure. 16 of 19 participants had trained with a split routine before the trial. | Methods, Training Procedures |
| Ochi et al. (2018) | 1 vs 3 | 12 repetitions at a fixed 67% 1RM (leg extension; T1 6 sets x1/wk, T3 2 sets x3/wk); not to failure: the authors state that they believe subjects did not train to failure, and the Methods allowed a rest when repetitions could not be completed. | Methods, Training Programs |
| Thomas & Burns (2016) | 1 vs 3 | 8–12 repetitions (≈75–85% 1RM); all sets performed to momentary muscular failure. | Methods, Protocol |
| Neves et al. (2022) | 1 vs 3 | Linear periodization 12 to 10 to 8 RM, RM-based in both limbs in the sets-equated condition used here; in the trial's volume-load-equated condition (sensitivity analysis) the higher-frequency limb was held to the lower-frequency limb's mean load and did not always reach momentary failure (explicitly noted). | Methods, training protocol |
| Calzavara & Barroso (2026) | 1 vs 3 | 3 weekly sets per exercise at 80% of pretest 1RM to concentric failure. | Abstract; Methods, Resistance Training Program |
| Candow & Burke (2007) | 2 vs 3 | 10 repetitions at 60–90% 1RM; each set to muscular fatigue at a self-selected load (load adjusted so fatigue occurred near the end of each set); momentary failure not stated. | Methods, RT Program |
| Lasevicius et al. (2019) | 2 vs 3 | 8–12 repetitions; all sets to the point of momentary concentric muscular failure (10RM-based load, ±2.5% progression). | Methods, RT procedures |
| Pina et al. (2020) | 2 vs 3 | 10–15 RM per set (seated calf 15–20 RM); RM-based loading, volitional failure not explicitly stated. | Methods, RT Program |
| Grzyb et al. (2022) | 2 vs 3 | 20–30 repetitions per set to volitional fatigue (defined as inability to complete the concentric phase); light-load, high-repetition. | Methods, HRRT Program |
| Brigatto et al. (2022) | 2 vs 3 | 8–12 RM; all sets to momentary concentric muscular failure (reported RPE/RIR of 9.5–10 for all sets). | Methods, training protocol |

*Contrast is taken from Table 1 of the main text. All sixteen prescriptions were extracted directly from each trial's Methods. Eleven trials prescribed working sets to muscular failure or volitional fatigue (Gentil 2015, Gentil 2018, Brigatto 2019, McLester 2000, Schoenfeld 2015, Thomas & Burns 2016, Calzavara & Barroso 2026, Candow & Burke 2007, Lasevicius 2019, Grzyb 2022, Brigatto 2022); four used repetition-maximum or percentage-based loading without confirming failure (Yue 2018, Prieto González 2020, Pina 2020, Neves 2022); and one used a fixed submaximal load, not to failure (Ochi 2018). The two Brigatto trials reported effort on a repetitions-in-reserve scale (RPE/RIR 9.5–10). In Schoenfeld 2015 and Brigatto 2019 the higher-frequency schedule was the less accustomed one; in Gentil 2018 the once-weekly schedule was.*

**Table S2. Records excluded at eligibility assessment, with reasons**

| Reason | Study | Assessed on | Detail |
|---|---|---|---|
| Weekly volume not equated | Arhab et al. (2023) | Abstract | Multicomponent clinical exercise program |
| Weekly volume not equated | Benton et al. (2011) | Full text | Body composition by plethysmography |
| Weekly volume not equated | Calaway et al. (2025) | Abstract | Same per-session velocity-loss prescription at 2 versus 3 days per week |
| Weekly volume not equated | Carneiro et al. (2015) | Abstract | 1 set per exercise per session in both arms; flexibility outcome |
| Weekly volume not equated | Cavalcante et al. (2018) | Abstract | Same session prescription |
| Weekly volume not equated | Cigerci & Genc (2020) | Full text | 3 sets per session in both arms |
| Weekly volume not equated | do Nascimento et al. (2022) | Abstract | Same session prescription; detraining and retraining |
| Weekly volume not equated | Dos Santos et al. (2024) | Full text | 1 set per exercise per session in both arms |
| Weekly volume not equated | Elmahgoub et al. (2011) | Abstract | Combined aerobic and resistance training, weekly volume not equated; full text not obtained |
| Weekly volume not equated | Elshakankiry et al. (2026) | Abstract | 1+1 versus 2+2 versus 3+3 sessions; combined training |
| Weekly volume not equated | Ferrari et al. (2013) | Abstract | Identical volume per session; concurrent training |
| Weekly volume not equated | Ferrari et al. (2016) | Abstract | Identical session volume; concurrent training |
| Weekly volume not equated | Fisher et al. (2013) | Abstract | 1+1 versus 2+2 versus 3+3 sessions; combined training |
| Weekly volume not equated | Geneen et al. (2022) | Abstract | Stated by authors |
| Weekly volume not equated | Ihalainen et al. (2019) | Abstract | Frequency increased volume, stated |
| Weekly volume not equated | Kawama et al. (2024) | Full text | 600 versus 900 repetitions |
| Weekly volume not equated | Martínez-Rodríguez et al. (2018) | Abstract | Body-weight circuit training, weekly volume not equated |
| Weekly volume not equated | Medeiros et al. (2020) | Full text | Same sets per session |
| Weekly volume not equated | Murlasits et al. (2012) | Full text | 3 sets per session in both arms |
| Weekly volume not equated | Pina et al. (2019) | Abstract | Same session prescription |
| Weekly volume not equated | Pina et al. (2020; a different trial from the included Pina et al., 2020) | Abstract | Same session prescription |
| Weekly volume not equated | Refsnes et al. (2025) | Abstract | Equal session doses, 1/2/3 per week |
| Weekly volume not equated | Reis et al. (1995) | Abstract | Same 3x12 session prescription; frequency varied by cycle phase, not fixed at 1/2/3 |

| Reason | Study | Assessed on | Detail |
|---|---|---|---|
| Weekly volume not equated | Sakamaki-Sunaga et al. (2016) | Abstract | 3 sets per session in both arms |
| Weekly volume not equated | Taaffe et al. (1999) | Abstract | 3 sets per session in all arms |
| Weekly volume not equated | Turpela et al. (2017) | Abstract | Same session prescription, 1/2/3 sessions |
| Weekly volume not equated | Van Vossel et al. (2023) | Full text | Higher volume in 3x limb, stated |
| Weekly volume not equated | Walker et al. (2018) | Abstract | Reduced-training phase only (1 versus 2 after common training); weekly volume not equated |
| Frequency outside 1 to 3 sessions per muscle per week | Arazi et al. (2021) | Abstract | 2 versus 4 |
| Frequency outside 1 to 3 sessions per muscle per week | Bartolomei et al. (2021) | Full text | Full-body 4 versus split 1 per muscle as described by authors |
| Frequency outside 1 to 3 sessions per muscle per week | Boyes et al. (2017) | Abstract | 10 versus 3 per week; isometric handgrip |
| Frequency outside 1 to 3 sessions per muscle per week | Braz et al. (2022) | Abstract | 2 versus 4 |
| Frequency outside 1 to 3 sessions per muscle per week | Canton-Martínez et al. (2022) | Abstract | 3 versus 5; concurrent training |
| Frequency outside 1 to 3 sessions per muscle per week | Carneiro et al. (2024) | Abstract | 1 versus 5 per muscle |
| Frequency outside 1 to 3 sessions per muscle per week | Colquhoun et al. (2018) | Abstract | 3 versus 6 |
| Frequency outside 1 to 3 sessions per muscle per week | Damas et al. (2019) | Abstract | 5 versus 2 to 3 |
| Frequency outside 1 to 3 sessions per muscle per week | Evangelista et al. (2021) | Full text | 2 versus 4 per muscle |
| Frequency outside 1 to 3 sessions per muscle per week | Firoozi et al. (2020) | Abstract | 3 versus 4 |
| Frequency outside 1 to 3 sessions per muscle per week | Franco et al. (2021) | Abstract | 1 versus up to 5 per muscle |
| Frequency outside 1 to 3 sessions per muscle per week | Gomes et al. (2019) | Abstract | 1 versus 5 |
| Frequency outside 1 to 3 sessions per muscle per week | Hamarsland et al. (2022) | Abstract | 2 versus 4 |

| Reason | Study | Assessed on | Detail |
|---|---|---|---|
| Frequency outside 1 to 3 sessions per muscle per week | Hartman et al. (2007) | Abstract | Twice versus once daily; 3-week intervention |
| Frequency outside 1 to 3 sessions per muscle per week | Maroto-Izquierdo et al. (2026) | Abstract | 1 versus 5 sessions per week |
| Frequency outside 1 to 3 sessions per muscle per week | Pedersen et al. (2024) | Abstract | 1 versus 4 |
| Frequency outside 1 to 3 sessions per muscle per week | Rukbumrung et al. (2025) | Full text | 2 versus 3 versus 4 sessions per week; size by bioimpedance and circumferences |
| Frequency outside 1 to 3 sessions per muscle per week | Saric et al. (2019) | Abstract | 3 versus 6 |
| Frequency outside 1 to 3 sessions per muscle per week | Zaroni et al. (2019) | Abstract | 1 versus 5 per muscle |
| Trial registration without results | Trial registry record: Are shorter resistance training sessions better than longer? (2022) | Abstract | Per-muscle frequency identical (2 per week in both arms); bioimpedance |
| Trial registration without results | Trial registry record: Effect of formatted resistance training on muscle mass and function in the elderly (2022) | Abstract | Frequency outside 1 to 3 and weekly volume not equated |
| Trial registration without results | Trial registry record: Effects of different resistance training modalities on muscle strength gains and urinary titin concentration dynamics (2026) | Abstract | Ongoing, 1 versus 3; volume equation not stated; no results |
| Trial registration without results | Trial registry record: Effects of Exercise Frequency on Muscle Size and Strength in Healthy, Untrained Young Adults | Abstract | Frequency outside 1 to 3 (2 versus 10 per week); no results |
| Trial registration without results | Trial registry record: Exploring How Exercise Frequency Impacts Muscle Resistance and Balance in Institutionalized Seniors (2025) | Abstract | Volume equation not stated; no results |
| Trial registration without results | Trial registry record: Frequency of Aerobic/Resistance Training in Older Women | Abstract | Registration of Fisher et al. (2013); weekly volume not equated |
| Trial registration without results | Trial registry record: Get in Shape in the Team Research: porukalla Kuntoon Tutkimus | Abstract | Registration of the Turpela and Ihalainen trial; weekly volume not equated |

| Reason | Study | Assessed on | Detail |
|---|---|---|---|
| | (2015) | | |
| Trial registration without results | Trial registry record: Use of Exercise Snacks to Improve Upper Body Fitness (2026) | Abstract | Exercise-snack design, frequency outside 1 to 3; no results |
| No per-muscle frequency contrast | Calder et al. (1994) | Abstract | Per-muscle frequency identical (2 per week in both arms) |
| No per-muscle frequency contrast | Izquierdo et al. (2004) | Abstract | Not a resistance-training frequency comparison (resistance versus resistance + endurance) |
| No per-muscle frequency contrast | Pedersen et al. (2022) | Abstract | Per-muscle frequency identical (2 per week in both arms) |
| No per-muscle frequency contrast | Stasinaki et al. (2015) | Abstract | Not a frequency comparison (compound versus complex training, same frequency) |
| Frequency self-selected or set by attendance | Grannell et al. (2021) | Abstract | Observational; frequency self-reported, not assigned |
| Frequency self-selected or set by attendance | Hoffman et al. (1990) | Abstract | Non-randomized self-selected frequency (3 to 6 days); strength outcomes only |
| Frequency self-selected or set by attendance | Kemmler & Von Stengel (2013) | Abstract | Retrospective grouping by attendance; multicomponent exercise |
| Frequency self-selected or set by attendance | Orsatti et al. (2014) | Abstract | Non-randomized; groups formed by attended frequency |
| Muscle size by bioimpedance only | Cardoso et al. (2024) | Full text | Full body (3 per muscle) versus A/B split (1.5 per muscle on average), volume equated, 8 weeks; fat-free mass by bioimpedance only |
| Shorter than six weeks | Ribeiro et al. (2015) | Abstract | 4 weeks; 4 versus 6 sessions per week |
| Conference abstract, not a trial | Mikula et al. (2025) | Abstract | Meta-regression presented as a conference abstract |
| **From other sources** | | | |
| No per-muscle frequency contrast | Arazi & Asadi (2011) | Full text | Each exercise performed once weekly in every group (only the grouping of exercises into sessions changed); size outcomes limited to skinfold body fat and limb circumferences |
| Weekly volume not equated | Baxter et al. (2024) | Abstract | Same 12-minute session once versus twice weekly |
| Weekly volume not equated | Schoenfeld et al. (2014) | Review table | Volume load equated across different loading schemes (3 |

| Reason | Study | Assessed on | Detail |
|---|---|---|---|
| | | | x 10RM versus 7 x 3RM), so frequency is confounded with load and sets are not matched |
| Shorter than six weeks | Crewther et al. (2016) | Review table | Four weeks per condition (crossover) |
| Frequency outside 1 to 3 sessions per muscle per week | Hunter (1985) | Review table | Three versus four sessions per week |

*The 66 records excluded at eligibility assessment from the database searches (Figure 1), grouped by the reason on which the exclusion rests, followed by the five reports from other sources that were excluded; for three of these the design was taken from the table of included studies of the review that cited them. Records were assessed on the full text where the abstract left any criterion open, and on the abstract where it stated the failing design feature explicitly. Where a record fails several criteria, the first listed is the one recorded. Cardoso et al. (2024) is the one trial excluded only for its muscle-size measure; its split arm also trained each muscle 1.5 times per week on average. Calaway et al. (2025) and Elmahgoub et al. (2011) were excluded on the abstract without the full text, which could not be obtained. Tavares et al. (2017), analyzed in sensitivity analysis only, is described in Section S2. The decision and reason for every record screened are in the screening log deposited on OSF.*

#### Table S3. Records identified by database

| Source | Platform | Records identified | Date searched |
|---|---|---|---|
| MEDLINE | PubMed | 709 | 6 Oct 2026 |
| Scopus | Elsevier | 1,178 | 6 Oct 2026 |
| Web of Science Core Collection | Clarivate | 567 | 6 Oct 2026 |
| SPORTDiscus with Full Text | EBSCO | 378 | 6 Oct 2026 |
| CENTRAL | Ovid EBM Reviews | 773 | 6 Oct 2026 |
| Total (with duplicates) | | 3,605 | |
| Duplicates removed | | 1,670 | |
| Records screened on title and abstract | | 1,935 | |
| Records assessed for eligibility | | 79 | |
| Reports assessed from other sources | | 10 | |
| Trials included (pooled / sensitivity only) | | 17 (16 / 1) | |

*Counts after each database's limits (S1). The per-source figures sum to the total identified (709 + 1,178 + 567 + 378 + 773 = 3,605). Of the 1,935 unique records, CENTRAL contributed 269 that no other database returned, all of which were excluded. Of the 709 PubMed records, 692 had entered PubMed by 15 April 2026, the end of the original search period.*

### Table S4. Moderator values and counting rule

| Trial | Lower-frequency arm | Measured muscle(s) | Sets per session | Sets per week | Mean repetitions | Relative load (% 1RM) |
|---|---|---|---|---|---|---|
| Gentil et al. (2015) | 1x/wk | Elbow flexors (lat pull-down, seated row and two curls, 3 sets each) | 12 | 12 | 10 | 75 |
| Gentil et al. (2018) | 1x/wk | Elbow flexors (same exercises as Gentil et al., 2015) | 12 | 12 | 10 | 75 |
| Yue et al. (2018) | 1x/wk (lower body) | Vastus medialis (parallel squat and deadlift in routine 2, 4 sets each) | 8 | 8 | 10 | 75 |
| Prieto González et al. (2020) | 1x/wk (split) | Whole body (skinfold); per major muscle group | 6 | 6 | 9 | 70 |
| Brigatto et al. (2019) | 1x/wk | Elbow flexors, triceps, vastus lateralis, anterior quadriceps | 16 | 16 | 10 | 75 |
| McLester et al. (2000) | 1x/wk | Whole body (skinfold); one exercise per muscle group | 3 | 3 | 6.5 | 82 |
| Schoenfeld et al. (2015) | 1x/wk (split) | Vastus lateralis (elbow flexors and extensors 6 direct sets) | 9 | 9 | 10 | 75 |
| Ochi et al. (2018) | 1x/wk | Quadriceps (leg extension only) | 6 | 6 | 12 | 67 |
| Thomas & Burns (2016) | 1x/wk | Whole body (DXA); per muscle group | 9 | 9 | 10 | 75 |
| Neves et al. (2022) | 1x/wk limb | Quadriceps (unilateral leg press only) | 9 | 9 | 10 | 75 |
| Calzavara & Barroso (2026) | 1x/wk | Quadriceps (back squat, leg press and leg extension, 3 sets each) | 9 | 9 | 8 | 80 |
| Candow & Burke (2007) | 2x/wk | Whole body (DXA); two exercises for | 6 | 12 | 10 | 75 |

| Trial | Lower-frequency arm | Measured muscle(s) | Sets per session | Sets per week | Mean repetitions | Relative load (% 1RM) |
|---|---|---|---|---|---|---|
| | | the major muscle groups | | | | |
| Lasevicius et al. (2019) | 2x/wk (split) | Quadriceps (squat, leg press, leg extension; elbow flexors and extensors 12 each) | 18 | 36 | 10 | 75 |
| Pina et al. (2020) | 2x/wk | Whole body (DXA); one exercise per muscle group | 3 | 6 | 12.5 | 68 |
| Grzyb et al. (2022) | 2x/wk | Elbow and knee flexors and extensors, one exercise each | 3 | 6 | 25 | 58 |
| Brigatto et al. (2022) | 2x/wk | Elbow flexors, triceps, vastus lateralis, anterior quadriceps | 9 | 18 | 10 | 75 |

*Counting rule: for each measured muscle, every set in the lower-frequency arm's session of an exercise in which that muscle is a prime mover is counted, multi-joint and single-joint alike (the rule under which Brigatto et al., 2019, describe 16 sets per muscle group as 8 multi-joint and 8 single-joint sets); where sites differ in their count, the quadriceps site is used where measured and otherwise the direct count for the measured muscle; for whole-body indirect measures, the count for a major muscle group. Weekly sets are the per-session count multiplied by the sessions per week for that muscle. Mean repetitions are the midpoint of the prescribed range; relative load is the stated percentage of one-repetition maximum or the approximate equivalent of the prescribed repetition-maximum range. Under the counting used in the submitted version, which mixed rules (sets per exercise for Lasevicius, 6 and 12; Schoenfeld 7; Candow 3 and 6; direct sets only for Gentil 2015, 6), the per-session slope was +0.008 g per set (p = 0.76) and the weekly slope +0.014 (p = 0.56); under the rule above they are +0.024 (p = 0.24) and +0.016 (p = 0.15). Neither coding gives a slope the regressions could have detected.*

### Table S5. Brigatto et al. (2022): composite effect size under alternative dispersion sources

| Source of change-score dispersion | Site d (MTBB, MTTB, MTVL, MTAQ) | Composite g [95% CI] | Overall pooled g [95% CI] |
|---|---|---|---|
| Trial-reported between-group effect sizes of the changes (used) | -0.31, -0.61, +0.60, -0.77 | -0.26 [-1.01, 0.49] | -0.06 [-0.24, 0.13] |
| Standard deviations implied by the group-by-time F statistics | -0.31, -0.61, +1.04, -0.77 | -0.16 [-0.91, 0.60] | -0.05 [-0.24, 0.13] |
| Standard deviations | -0.54, -1.40, +2.10, -1.46 | -0.31 [-1.14, 0.52] | -0.06 [-0.24, 0.13] |

| Source of change-score dispersion | Site d (MTBB, MTTB, MTVL, MTAQ) | Composite g [95% CI] | Overall pooled g [95% CI] |
|---|---|---|---|
| recovered from the printed 95% CIs of the mean change | | | |
| Pre and post standard deviations with r = 0.7 (imputation) | -0.22, -0.24, +0.36, -0.27 | -0.09 [-0.83, 0.65] | -0.05 [-0.23, 0.14] |

*Negative favors the higher frequency; n = 10 per group; composite with a within-trial correlation of 0.6. The trial-reported effect sizes agree with the standard deviations implied by its group-by-time F statistics at three of the four sites (the exception is the vastus lateralis, where the printed F and p values are themselves inconsistent), whereas the standard deviations implied by its printed confidence intervals are 1.7 to 3.5 times smaller than those implied by the reported effect sizes and would imply between-group differences the trial's own tests did not find. The reported effect sizes were therefore used.*

#### Table S6. Confidence intervals for the overall estimate and each contrast under alternative conventions

| Pool | k | g | Wald 95% CI (reported) | HKSJ, truncated at q = 1 | HKSJ, unmodified | Q (df), p |
|---|---|---|---|---|---|---|
| Overall | 16 | -0.06 | -0.24 to 0.13 | -0.26 to 0.14 | -0.21 to 0.09 | 8.45 (15), 0.90 |
| 1 vs 2 | 5 | +0.07 | -0.29 to 0.43 | -0.44 to 0.58 | -0.33 to 0.47 | 2.50 (4), 0.64 |
| 1 vs 3 | 6 | -0.23 | -0.57 to 0.10 | -0.67 to 0.21 | -0.48 to 0.01 | 1.55 (5), 0.91 |
| 2 vs 3 | 5 | -0.01 | -0.29 to 0.27 | -0.41 to 0.38 | -0.34 to 0.32 | 2.79 (4), 0.59 |
| Overall, omitting Lasevicius et al. (2019) | 15 | -0.10 | -0.30 to 0.09 | -0.32 to 0.11 | -0.24 to 0.04 | 5.99 (14), 0.97 |

*The unmodified HKSJ interval uses the observed dispersion of the trials about the pooled estimate as the scaling factor; when that dispersion is below its expectation (Q below its degrees of freedom), the interval is narrower than the Wald interval. In the submitted twelve-trial analysis, the unmodified interval excluded zero with Lasevicius et al. (2019) omitted; in the sixteen-trial analysis it does not. The truncated version (scaling factor not below one) with t on k - 1 degrees of freedom is wider than the Wald interval at every k.*

### Supplementary Figures

#### Figure S2. Per-trial effects by frequency contrast (paneled forest plot)

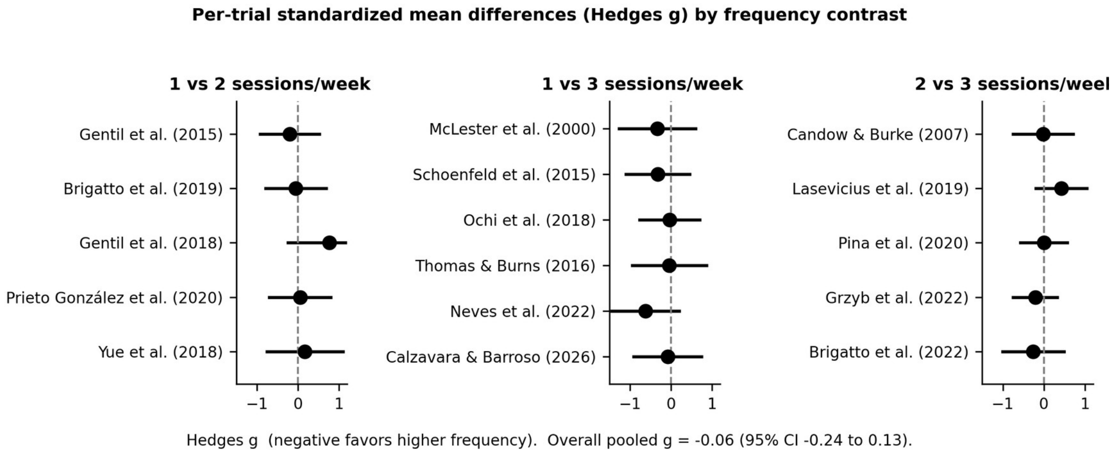


***Figure S2.*** *Per-trial change-score standardized mean differences (Hedges g, with 95% confidence intervals) for the sixteen pooled trials, arranged in panels by frequency contrast (1 vs 2, 1 vs 3, and 2 vs 3 sessions per muscle per week). Negative values favor the higher-frequency condition. The overall pooled estimate is g = -0.06 (95% CI -0.24 to 0.13). This is a paneled presentation of the main-text forest plot (Figure 2).*
